# Association between the antibiotics use and recurrence in patients with resected non-metastatic colorectal cancer: EVADER-1, a nation-wide pharmaco-epidemiologic study

**DOI:** 10.1101/2023.09.23.23296001

**Authors:** Marc Hilmi, Ines Khati, Anthony Turpin, Antoine Andremont, Charles Burdet, Nathalie Grall, Joana Vidal, Philippe-Jean Bousquet, Benoît Rousseau, Christine Le Bihan-Benjamin

## Abstract

**Importance:** The impact of antibiotics (ATBs) on the risk of colorectal cancer (CRC) recurrence after curative resection remains unknown.

**Objective:** We aimed to evaluate the effect of the type and the class of ATBs on recurrence in patients with resected non-metastatic CRC.

**Design:** Our cohort study included patients between 01/2012 and 12/2014. Each CRC patient was followed up to 3 years after surgical resection.

**Setting:** This study was conducted using extracted data from the French cancer cohort set up based on the National Health Data System by the French National Cancer Institute.

**Participants:** All newly diagnosed non-metastatic CRC patients resected were included. We excluded patients not affiliated to the major health insurance scheme in France (n=16 857) and with immunosuppression (n=32,359).

**Exposures:** The perioperative ATB intake (from 6 months before surgery until 1 year after) was classified according to the class, the period of use (pre- vs post-resection), the disease stage (localized and locally advanced), and the primary tumor location (colon and rectum/junction).

**Main Outcome and Measure:** The primary endpoint was the 3-year disease-free survival (DFS). The impact of ATB was assessed using time-dependent multivariate Cox models.

**Results:** A total of 35,496 CRC patients were included. Seventy-nine percent of patients had at least one ATB intake. Outpatient ATB intake after surgery was associated with unfavorable 3-year DFS mostly in patients with locally advanced stage and during chemotherapy (HR ranging from 1.22 to 1.41, P<0.0001), while no excess of mortality was observed in patients receiving ATBs as outpatient. The ATBs associated with decreased 3-year DFS were cephalosporins, streptogramins, quinolones, penicillin A with beta-lactamase inhibitors, and antifungals with differential effects according to the primary tumor location and disease stage.

**Conclusion and Relevance:** These findings suggest that ATBs modulate the risk of recurrence after early CRC resection with a differential impact of the ATB classes depending on disease stage and tumor site. This study also gives important clues on how ATBs may modulate the efficacy of cancer treatments. Ultimately, EVADER-1 will pave the way for therapeutic interventions targeting the microbiome aiming to improve cancer outcome.

**Key points:** *Question:* What is the impact of antibiotics on the risk of colorectal cancer recurrence after curative intent resection?

*Findings:* In this homogenous cohort of patients with resected colorectal cancer, the antibiotic intake had a differential effect depending on the primary location, the disease stage, the timing of intake relative to chemotherapy and the type of antibiotics.

*Meaning:* The interaction of antibiotics with disease-free survival encourages limiting long-term/multiple antibiotic exposure and supports therapeutic interventions targeting the microbiome to improve cancer outcome.

## INTRODUCTION

Colorectal cancer (CRC) accounts for 1.8 million new cases per year worldwide and is the third cause of cancer mortality in Europe and the United States.^1^ Despite the optimal management at the non-metastatic stage, about 25% of patients will recur within the first 3 years after cancer surgery.^2^ Thus, identification of new prognostic factors and therapeutic strategies is crucially needed.

Antibiotics (ATBs) durably modify the digestive microbiota by decreasing its diversity, which full reconstitution takes several months.^3^ Dysbiosis, defined as an imbalance of the microbiota, promotes several diseases including cancer.^4^ Epidemiological cohort studies showed a positive association between ATBs consumption and the incidence of CRC (hazard ratio [HR]=2.01, P<0.05),^5^ and digestive adenomas (HR=1.36, P<0.05).^6^ A Swedish case-control registry study of more than 40,000 CRC patients strengthened the level of evidence on the effect of the ATB use by showing an increased risk of proximal colon cancer (HR=1.17) and an inverse effect on rectum cancer (HR=0.84).^7^

While the association between ATB and the incidence of CRC has been suggested,^5^ the impact of an ATB consumption on the recurrence of non-metastatic CRC after resection is unknown. The 3-year disease-free survival (DFS) is a validated criterion for adjuvant studies^8^ and an accepted surrogate for overall survival in CRC. Our hypothesis was that perioperative ATB consumption may modulate the 3-year DFS of patients with non-metastatic CRC. In this extensive pharmaco-epidemiologic study based on comprehensive real-world data, we analyzed the effect of the type and class of ATB on recurrence in patients with resected non-metastatic CRC.

## MATERIALS AND METHODS

### Data sources

This prospective study was conducted using extracted data from the French cancer cohort set up on the basis of the National Health Data System by the French National Cancer Institute^9,10^. The cohort included all people diagnosed, treated, or followed up for a cancer in France since 2010.^11^ Data on ATBs use related to hospitalization care or outpatient care were extracted. The French cancer cohort protocol was approved by a national committee and authorized by the French Data Protection Agency (CNIL, number 2019-082), and was conducted in accordance with the Declaration of Helsinki.

### Selection criteria

All people aged 18 or older with incident of non-metastatic CRC resected between January 2012 and December 2014 were selected. Cases with a previous history of cancer (2010-2011) or long-term disease for cancer (diagnosed before 2012) were excluded. In addition, individuals with a concomitant cancer or metastatic CRC diagnosed simultaneously or within 3 months of the initial diagnosis, with immunosuppression (including HIV, immunosuppressant intake of >1 month during the study duration, transplant, and organ transplantation codes), living abroad, not affiliated to the major health insurance scheme in France, with an unknown ICD-10 codes of surgical resection or double tumor location (colon and junction), with a >3-month delay between the date of the first hospitalization after cancer diagnosis/the last neoadjuvant treatment and the date of surgical resection, were also excluded. To obtain a homogeneous population with the same study duration, each CRC patient was followed up to 3 years after surgical resection.

### Measurement of the ATB exposure

All reimbursements for ATBs were extracted for the 6-month prior surgery and 1-year after. We assumed that each pill delivered was taken by the patient. The daily dose of each ATB was inferred from the number of days of treatment reimbursed. We then considered that each ATB had an effect on the microbiota during the days of intake and the 30-day follow-up period to consider the potential overlap with anticancer treatment.

Because drugs dispensed during hospitalization were not directly traceable in the cohort, the consumption of ATB was evaluated by the presence of a diagnostic code (ICD-10) for infectious diseases. To isolate post-operative infections, hospital infections occurring after surgery were divided into two categories: occurring within 30 days and more than 30 days after surgery.

For outpatient consumption of ATB, an aggregation of medications was carried out according to their spectrum by two double-blinded infectiologists (Supplementary Table 1). Eight specific categories (azole antifungals, third generation cephalosporins [C3G]), macrolides, metronidazole-based ATB, penicillin A, penicillin A including beta-lactamase inhibitors, quinolones, streptogramins) and three non-specific categories (other antifungals, other beta-lactams, other ATB) of ATBs were defined.

### Covariates

Several potentially predictive factors for survival were identified or reconstructed from the available data: sex, age groups (18-49 years vs 50-69 years vs 70-79 years vs ≥ 80 years), Charlson comorbidity index through ICD-10 codes and specific acts and drugs^12^ for predicting non-cancer death (none vs 1-2 vs ≥3), nutritional status during the surgical resection (malnutrition vs no malnutrition) based on the ICD-10 codes for mild, moderate, or severe protein-based malnutrition (code ICD-10 E43, E44), the length of intensive care unit stay after surgery (no admission or <2 days vs 2-7 days vs >7 days).

The precise time-point of the ATB use to account for a potential time-dependent bias. If recurrence developed within the first year after resection, only the use of ATB before recurrence was considered in the model.

### Identification of recurrences

The occurrence of an ICD-10 diagnostic code for metastases, a palliative care code, cancer-related surgical resection following the first one, or treatment with chemotherapy or radiotherapy starting >3 months after the surgical resection was regarded as an indicator of recurrence through an algorithm applied to the French nation-wide database of cancer patients.

### Statistical analysis

The 3-year DFS was defined as the time between surgery and the date of the first documented recurrence, death, or the end of 3-year follow-up, whichever occurred first. The DFS curves were modelled using the Cox proportional hazards model to study the association between the ATB intake adjusted for all covariates and survival rates.

The survival curves were represented using the Kaplan-Meier method and compared with the log-rank test. Hazard ratios (HR) were estimated by the Cox proportional hazards regression model. Given the different prognoses in terms of recurrence and survival, the analyses were stratified according to four groups. These groups were constituted based on coding elements (diagnosis and treatment): localized colon cancer (surgical resection only), locally advanced colon cancer (surgical resection with neoadjuvant or/and adjuvant chemotherapy), localized rectum/junction cancer (surgical resection only), and locally advanced rectum/junction cancer (surgical resection and treatment with chemotherapy or/and radiotherapy).

Antibiotic exposure before T0 (surgery) was considered a fixed variable, while after T0 a time-dependent variable in a single change Cox model.^13,14^ The temporality relative to chemotherapy of antibiotic exposure after T0 was considered (during chemotherapy/not during chemotherapy). Two multivariate models were carried out on these four sub-populations: i/ considering the ATBs use in hospital and/or outpatient care, and ii/ considering single consumption by the ATBs categories. Specific multivariate models were performed on the population treated with chemotherapy to account for the use of ATBs depending on their timing of intake during or outside chemotherapy.

As the ATB hospital intake was more associated with negative prognostics factors and as ATBs administered during hospital stays are not recorded in the Cancer Cohort, the sensitivity analysis was performed excluding patients with infection during hospitalization to measure the outpatient ATB intake impact and to test the robustness of results. Associations of ATBs with the 3-year DFS were classified into 3 categories: 1) probable, if there was a significant association in the global and sensitivity analysis; 2) possible, if there was only a significant association in the sensitivity analysis, and 3) likely not present, if there was no significant association in both analysis or there was a significant association only in the global analysis. All statistical analyses were performed on WPS Analytics 4.0.

## RESULTS

### Patient population

A total of 121,532 patients were diagnosed with CRC between January 2012 and December 2014 in France (Figure 1). After excluding patients with synchronous metastases (32,359 [26.6%]) and those who did not meet the eligibility criteria, a total of 35,496 patients with resection of non-metastatic CRC were considered for the study (Figure 1). Colon cancer was the most prevalent primitive location (23,540 [66.3%] with 17,735 [75.3%] localized and 5,805 [24.7%] locally advanced). Patients with rectum/junction cancer (11,959 [33.7%]) had more frequently and locally advanced stage (6,859 [57.3%]).

**Figure 1.**
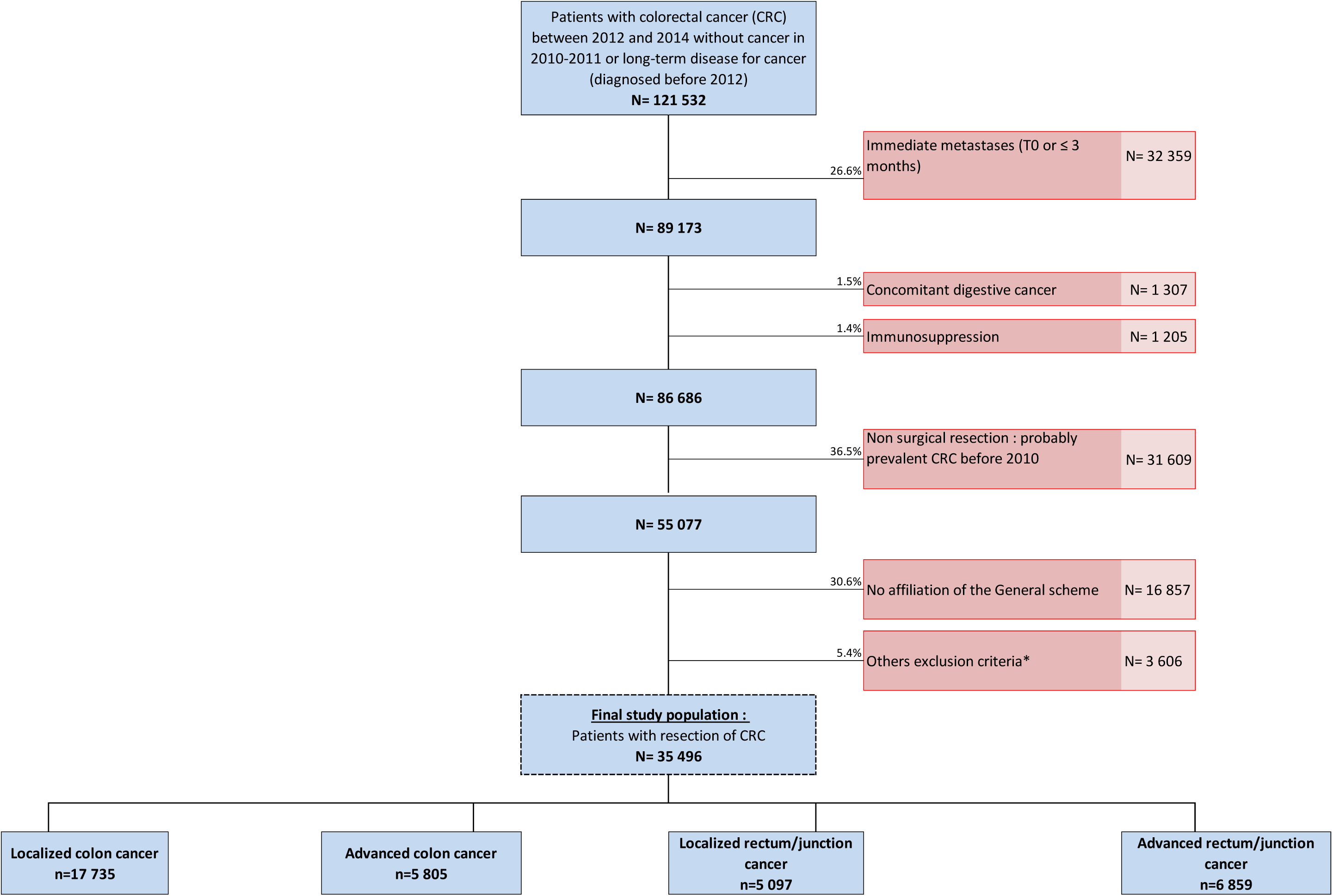
CONSORT-like workflow diagrams for the study cohort. * Living abroad, delay > 3 months between cancer diagnosis and the date of surgical resection or between the date of the last neoadjuvant treatment and the date of surgical resection, delay > 180 days between surgical resection and adjuvant chemotherapy, or unknown ICD-10 codes of surgical resection or double localization (colon and junction).

The demographic and clinical characteristics of the study cohort are represented in Table 1. Among all patients included, 52.3% were male (n=18,560) with the median age of 70 years, 12% were malnourished (n=4,250), and 64% had no comorbidities according to Charlson score (n=22,712). Of the patients with colon cancer, 23.7% received adjuvant chemotherapy (n=5,567) and 53.3% of those with rectum/junction cancer received neoadjuvant or adjuvant chemotherapy (n=6,368). After the 3 years of follow-up, 29.4% of patients relapsed (n=10,440) and 8.6% died (n=3,065).

**Table 1.**
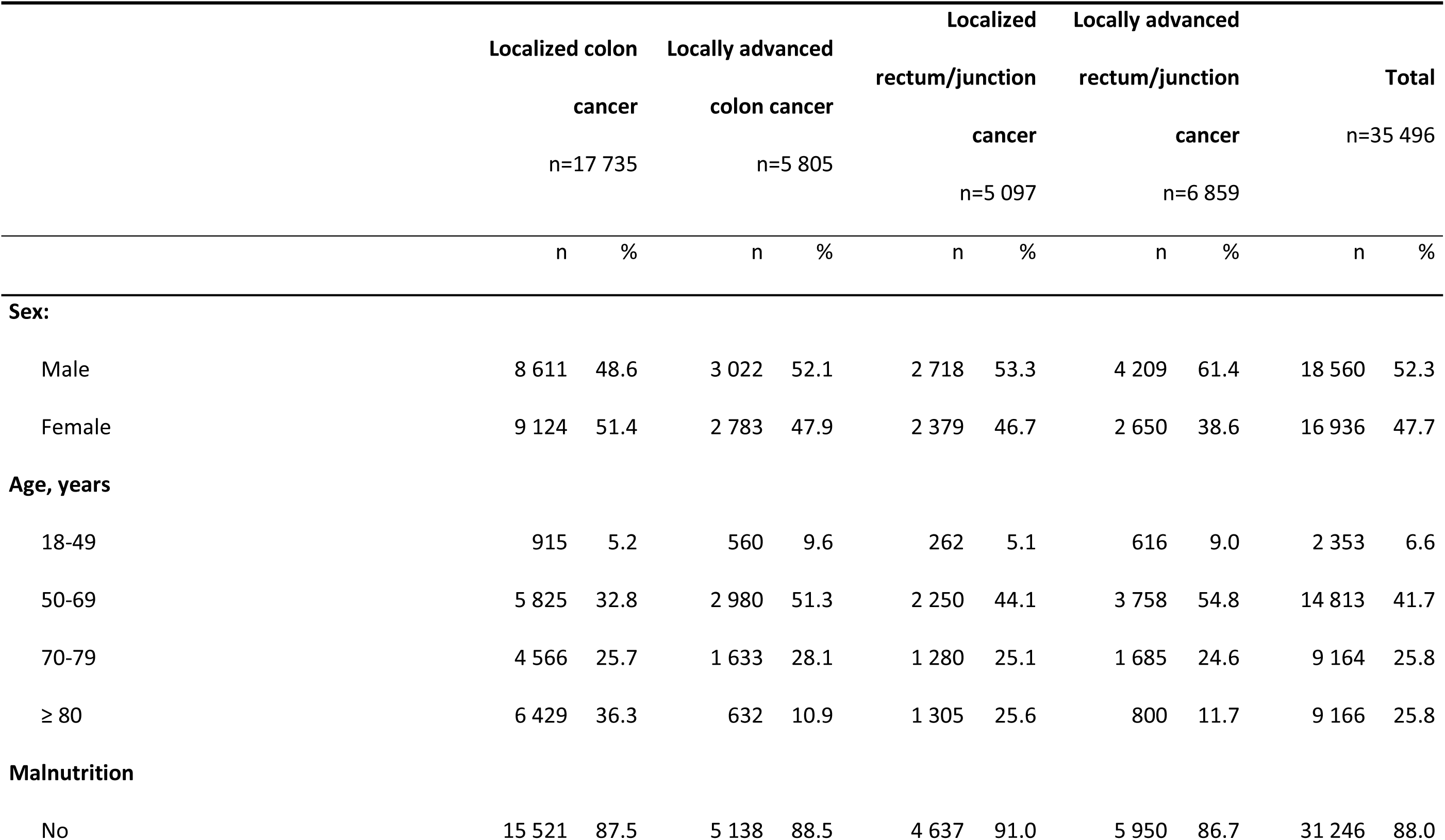
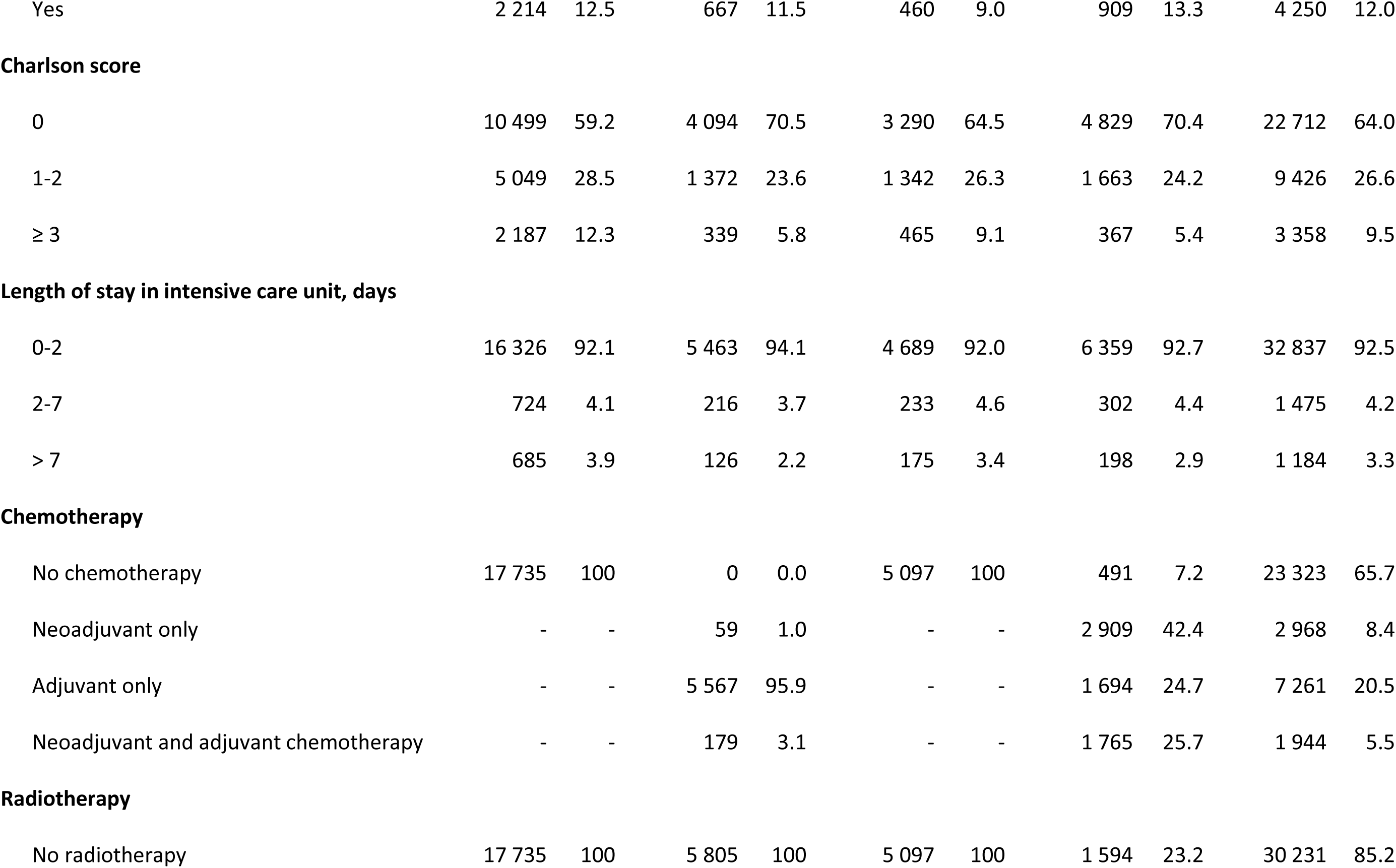
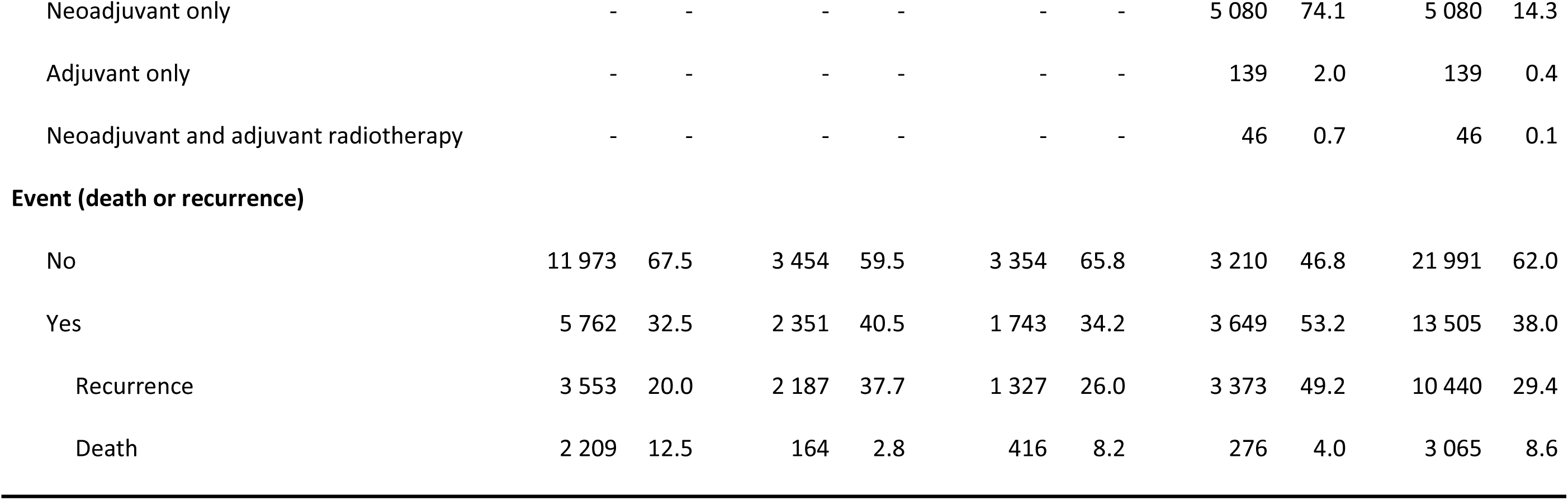
Clinical characteristics of patients with resection of colorectal cancer from the French nation-wide database (n= 36 105).

Across all included patients, 78.7% received at least one ATB in the perioperative setting (n=27,919) and were exposed mainly to quinolones (8,914 [25.1%], penicillin A (8,345, [23.5%]) and penicillin A combined with beta-lactamase inhibitor (8,413 [23.7%]) (Table 2), or to ATBs combinations (13,669 [38.5%]) (Supplementary Table 2). Furthermore, 37.6% of patients received an ATB during hospitalization (n=13,326) (Table 2), mostly for digestive infections (41.8%) and post-surgical site infection (38.5%) during the cancer surgery stay (Supplementary Table 3). Supplementary Tables 4 and 5 detail the prevalence of the ATB exposure for each class during the pre- and post-resection periods, and the number of chemotherapy days with ATB-related impact on microbiota.

**Table 2.**
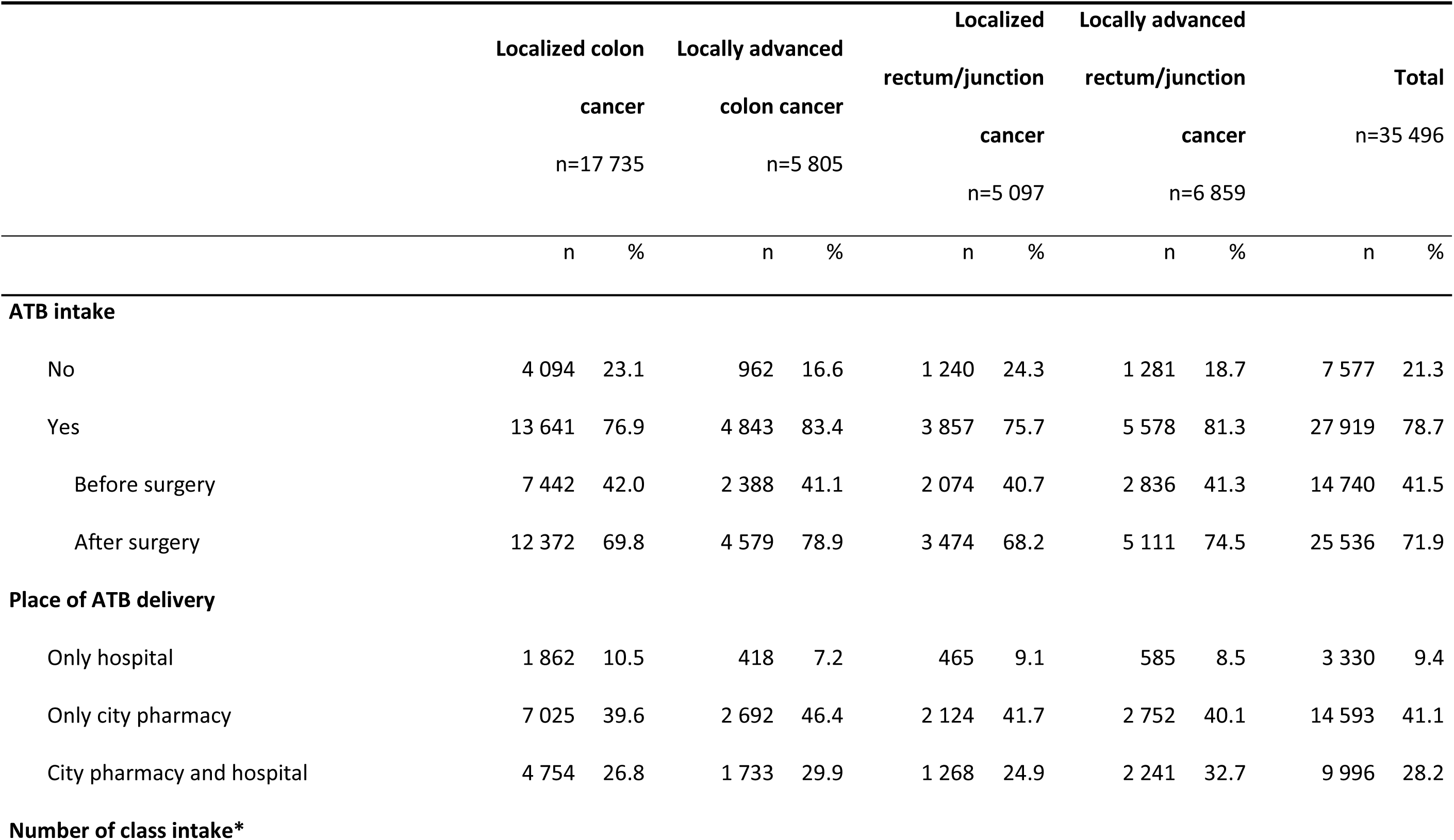
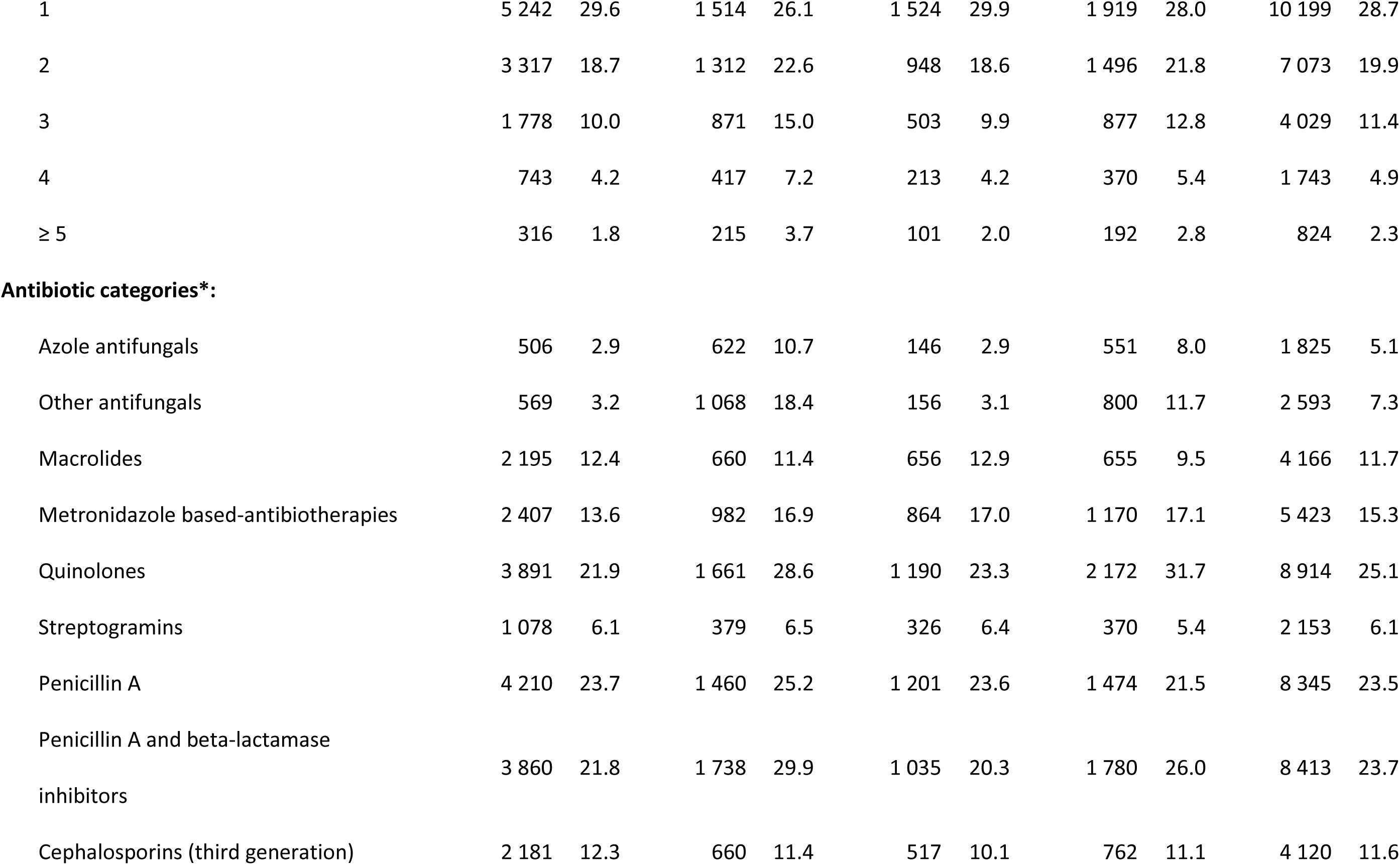
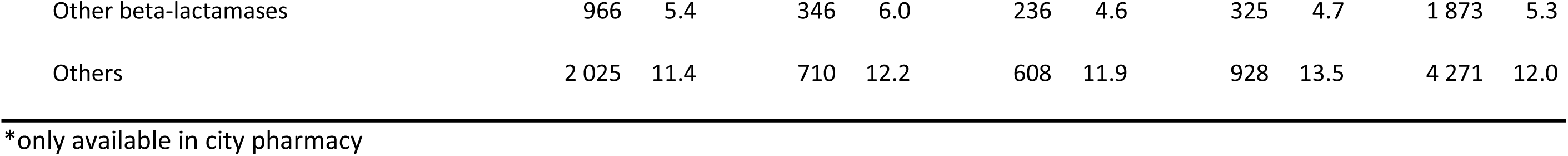
Antibiotics use in patients with resection of colorectal cancer included in the French nation-wide database (n= 35 496).

|  | Localized colon cancer<br>n=17 735 |  | Locally advanced colon cancer<br>n=5 805 |  | Localized rectum/junction cancer<br>n=5 097 |  | Locally advanced rectum/junction cancer<br>n=6 859 |  | Total<br>n=35 496 |  |
| --- | --- | --- | --- | --- | --- | --- | --- | --- | --- | --- |
|  | n | % | n | % | n | % | n | % | n | % |
| **ATB intake** |  |  |  |  |  |  |  |  |  |  |
| No | 4 094 | 23.1 | 962 | 16.6 | 1 240 | 24.3 | 1 281 | 18.7 | 7 577 | 21.3 |
| Yes | 13 641 | 76.9 | 4 843 | 83.4 | 3 857 | 75.7 | 5 578 | 81.3 | 27 919 | 78.7 |
| Before surgery | 7 442 | 42.0 | 2 388 | 41.1 | 2 074 | 40.7 | 2 836 | 41.3 | 14 740 | 41.5 |
| After surgery | 12 372 | 69.8 | 4 579 | 78.9 | 3 474 | 68.2 | 5 111 | 74.5 | 25 536 | 71.9 |
| **Place of ATB delivery** |  |  |  |  |  |  |  |  |  |  |
| Only hospital | 1 862 | 10.5 | 418 | 7.2 | 465 | 9.1 | 585 | 8.5 | 3 330 | 9.4 |
| Only city pharmacy | 7 025 | 39.6 | 2 692 | 46.4 | 2 124 | 41.7 | 2 752 | 40.1 | 14 593 | 41.1 |
| City pharmacy and hospital | 4 754 | 26.8 | 1 733 | 29.9 | 1 268 | 24.9 | 2 241 | 32.7 | 9 996 | 28.2 |
| **Number of class intake\*** |  |  |  |  |  |  |  |  |  |  |
| 1 | 5 242 | 29.6 | 1 514 | 26.1 | 1 524 | 29.9 | 1 919 | 28.0 | 10 199 | 28.7 |
| 2 | 3 317 | 18.7 | 1 312 | 22.6 | 948 | 18.6 | 1 496 | 21.8 | 7 073 | 19.9 |
| 3 | 1 778 | 10.0 | 871 | 15.0 | 503 | 9.9 | 877 | 12.8 | 4 029 | 11.4 |
| 4 | 743 | 4.2 | 417 | 7.2 | 213 | 4.2 | 370 | 5.4 | 1 743 | 4.9 |
| ≥ 5 | 316 | 1.8 | 215 | 3.7 | 101 | 2.0 | 192 | 2.8 | 824 | 2.3 |
| **Antibiotic categories\*:** |  |  |  |  |  |  |  |  |  |  |
| Azole antifungals | 506 | 2.9 | 622 | 10.7 | 146 | 2.9 | 551 | 8.0 | 1 825 | 5.1 |
| Other antifungals | 569 | 3.2 | 1 068 | 18.4 | 156 | 3.1 | 800 | 11.7 | 2 593 | 7.3 |
| Macrolides | 2 195 | 12.4 | 660 | 11.4 | 656 | 12.9 | 655 | 9.5 | 4 166 | 11.7 |
| Metronidazole based-antibiotherapies | 2 407 | 13.6 | 982 | 16.9 | 864 | 17.0 | 1 170 | 17.1 | 5 423 | 15.3 |
| Quinolones | 3 891 | 21.9 | 1 661 | 28.6 | 1 190 | 23.3 | 2 172 | 31.7 | 8 914 | 25.1 |
| Streptogramins | 1 078 | 6.1 | 379 | 6.5 | 326 | 6.4 | 370 | 5.4 | 2 153 | 6.1 |
| Penicillin A | 4 210 | 23.7 | 1 460 | 25.2 | 1 201 | 23.6 | 1 474 | 21.5 | 8 345 | 23.5 |
| Penicillin A and beta-lactamase inhibitors | 3 860 | 21.8 | 1 738 | 29.9 | 1 035 | 20.3 | 1 780 | 26.0 | 8 413 | 23.7 |
| Cephalosporins (third generation) | 2 181 | 12.3 | 660 | 11.4 | 517 | 10.1 | 762 | 11.1 | 4 120 | 11.6 |
| Other beta-lactamases | 966 | 5.4 | 346 | 6.0 | 236 | 4.6 | 325 | 4.7 | 1 873 | 5.3 |
| Others | 2 025 | 11.4 | 710 | 12.2 | 608 | 11.9 | 928 | 13.5 | 4 271 | 12.0 |

\*only available in city pharmacy

### Model construction

In univariate analysis, prognosis factors associated with a worse 3-year DFS, regardless of stage, included male gender, higher age, more comorbidities according to Charlson score, malnutrition at resection, and longer stay in intensive care unit. No significant differences in the mean number of chemotherapy cycles administered were observed according to the ATBs intake (Supplementary Figure 1).

To assess the overall impact of ATB intake on the risk or recurrence or death of resected non-metastatic CRC, four different models according to the primitive location and the disease stage were performed: localized colon cancer, locally advanced colon cancer, localized rectum/junction cancer, and locally advanced rectum/junction cancer. Univariate survival curves assessing the impact of the primitive location and stage are shown in the Supplementary Figure 2.

### Impact of antibiotic intake during hospitalization

The forest plots of main results are presented in Figure 2 and detailed results in Supplementary Table 6. Overall, in multivariate analyses we confirmed the prognosis association of most of the prementioned identified parameters in the model construction part.

**Figure 2.**
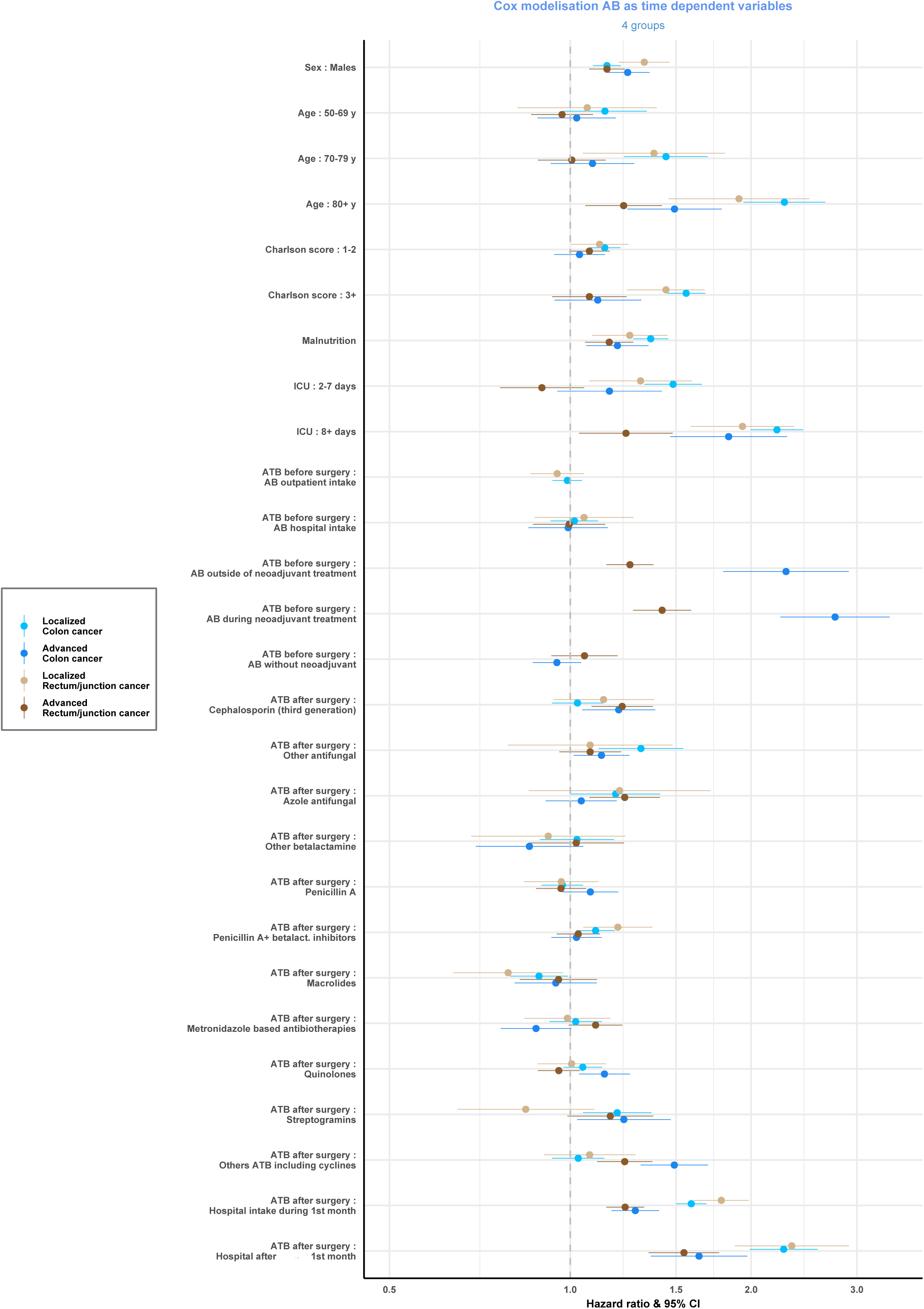
Multivariate analyses forest plots assessing the ATB intake impact on 3-year disease-free survival in resected colorectal cancer patients in four groups according to the cancer primitive location and stage. Multivariate analyses were adjusted on sex, age, Charlson score, malnutrition, and length of stay in intensive care unit. In case of chemotherapy exposure, the impact of ATB intake was assessed according to the timing of use relative to chemotherapy. Hazard ratios for each parameter and each model are available in Supplementary Table 6.

The ATBs hospital intake before surgery was not associated with the 3-year DFS (P>0.05), while ATBs intake after surgery was strongly associated with decreased 3-year DFS (P<0.0001) (Supplementary Table 6). The effect was more pronounced in case of localized stage and an intake more than 30 days after surgery (localized colon cancer: HR (≤30 days)=1.59 [1.50-1.69] and HR (>30 days)=2.27 [1.99-2.58]; locally advanced colon cancer: HR (≤30 days)=1.28 [1.17-1.41] and HR (>30 days)=1.64 [1.36-1.97]; localized rectum/junction cancer: HR (≤30 days)=1.78 [1.61-1.98] and HR (>30 days)=2.34 [1.88-2.91]; locally advanced rectum/junction cancer: HR (≤30 days)=1.23 [1.15-1.33] and HR (>30 days)=1.55 [1.35-1.77]).

To differentiate between a potential excess of deaths including the patients being hospitalized for infections, we assessed the difference between recurrence and death rates according to the place where patients received ATB (Supplementary Table 7). This analysis revealed a difference in the death rate of patients being hospitalized for infection (23.4%) compared to patients receiving or not ATB as outpatients (7.9% and 4.6%, respectively). Patients hospitalized for infection were more likely to be older (age ≥ 80 years old 30.8% vs 22.8%, P<0.0001), malnourished (17.7% vs 5.5%, P<0.0001), stayed longer in intensive care unit (more than 7 days 7.1% vs less than 7 days 1.1%, P<0.0001), and had more comorbidities (≥ 3 comorbidities 12.8 % vs < 3 comorbidities 7.4%, p<0.0001) (Supplementary Table 8).

Overall, the patients with infection during hospitalization seemed to have a worse 3-year DFS mainly because of an excess of deaths associated with worst prognostic factors. No statistical difference in the death rate of patients receiving ATBs as outpatients or not receiving it was observed.

### Impact of the outpatient ATBs intake

Summarized results of the association between the outpatient ATBs intake and 3-year DFS are presented in Table 3 and detailed results are presented in Supplementary Table 9. To avoid false discovery, we compared two models (global corresponding to all ATBs intake and sensitivity corresponding to a population excluding patients with infection during hospitalization). Concordant significant results between models were defined as probable and results only significant in sensitivity model as possible.

**Table 3.** Hazard ratios of multivariate models and the level of confidence of association between the outpatient antibiotic intake and 3-year disease-free-survival.

| Antibiotic exposure |  | Localization and stage |  |  |  |  |  |  |  |
| --- | --- | --- | --- | --- | --- | --- | --- | --- | --- |
|  |  | Localized colon cancer |  | Locally advanced colon cancer |  | Localized rectum junction cancer |  | Locally advanced rectum junction cancer |  |
|  |  | Global analysis | Sensitivity analysis | Global analysis | Sensitivity analysis | Global analysis | Sensitivity analysis | Global analysis | Sensitivity analysis |
| Pre resection | Without chemotherapy | 0.99 | 1.00 | 0.95 | 0.97 | 0.95 | 0.97 | 1.05 | 1.00 |
|  | Outside chemotherapy | NA | NA | 1.18 | 1.12 | NA | NA | 0.92 | 0.84* |
|  | During chemotherapy | NA | NA | 1.25 | 0.98 | NA | NA | 1.11* | 1.04 |
| Post resection | Without chemotherapy | 1.05 | 1.11* | NA | NA | 0.96 | 1.05 | NA | NA |
|  | Outside chemotherapy | NA | NA | 1.16* | 1.21* | NA | NA | 0.94 | 0.91 |
|  | During chemotherapy | NA | NA | 1.22*** | 1.32*** | NA | NA | 1.31*** | 1.41*** |
|  | C3G | 1.03 | 0.98 | 1.20** | 1.25* | 1.14 | 1.07 | 1.22*** | 1.21 |
|  | Antifungal-others | 1.31** | 1.19 | 1.13* | 1.22** | 1.08 | 1.21 | 1.09 | 1.06 |
|  | Antifungal-azoles | 1.19 | 1.45** | 1.04 | 0.93 | 1.21 | 1.10 | 1.24** | 1.18 |
|  | Betalactamins-others | 1.03 | 1.07 | 0.85 | 0.83 | 0.92 | 0.83 | 1.02 | 1.19 |
|  | Penicillin A | 0.97 | 0.98 | 1.08 | 1.09 | 0.97 | 1.04 | 0.97 | 1.02 |
|  | Penicillin A + inhibitors | 1.10** | 1.15* | 1.02 | 1.02 | 1.20** | 1.26* | 1.04 | 1.13 |
|  | Macrolides | 0.89* | 0.91 | 0.94 | 0.99 | 0.79* | 1.00 | 0.96 | 0.94 |
|  | Metronidazole | 1.02 | 1.03 | 0.88 | 1.04 | 0.99 | 0.96 | 1.10 | 1.20* |
|  | Quinolones | 1.05 | 1.16** | 1.14** | 1.16* | 1.01 | 1.03* | 0.96 | 1.01 |
|  | Streptogramins | 1.20** | 1.18 | 1.23* | 1.08 | 0.84 | 0.70 | 1.17 | 1.13 |
|  | Others including cyclins | 1.03 | 0.99 | 1.49*** | 1.55*** | 1.08 | 0.90 | 1.23*** | 1.40*** |

\*: $P \leq 0.05$ ; \*\*: $P \leq 0.01$ ; \*\*\*: $P \leq 0.001$ . Red : probable association; orange: possible association; blank : likely no association. Detailed results are presented in Supplementary Table 9.

A probable negative association between outpatient ATB intake and the 3-year DFS after resection was observed in locally advanced stages during chemotherapy for both location (HR ranging from 1.22 to 1.41, P<0.0001) and outside chemotherapy only in locally advanced colon cancer (HR (global analysis)=1.16 [1.02-1.33], HR (sensitivity analysis)=1.21 [1.01-1.45]; P=0.03)).

Regarding possible associations, outpatient ATBs intake before resection was associated with improved/3-year DFS in patients with locally advanced rectum/junction cancer receiving ATBs outside neoadjuvant treatment (HR=0.84 [0.72-0.98], P=0.03). A negative possible association between outpatient ATB intake and the 3-year DFS after resection was deemed possible for localized colon cancer: HR (sensitivity analysis)=1.11 [1.02-1.20], P=0.02.

### Differential impact of ATBs classes

Regarding the ATBs classes, all probable associations with the 3-year DFS were deleterious and differed according to the primary location and the disease stage (Table 3).

In the localized setting, a negative probable association between the “penicillin A + beta-lactamase inhibitors” class and the 3-year DFS was found for all primary location with HR ranging from 1.10 to 1.26. The association was possible for the class “antifungal-azoles” in localized colon cancer (HR=1.45 [1.12-1.87], P=0.005), for quinolones in localized colon cancer (HR=1.16 [1.04-1.30], P=0.009) and localized rectum/junction cancer (HR=1.26 [1.03-1.53], P=0.02).

In the locally advanced setting, negative probable associations were observed for C3G, quinolones, and "antifungal-others" classes in patients with locally advanced colon cancer with HR ranging from 1.13 to 1.25. A negative probable association between the class “others-including cyclins” and the 3-year DFS was observed for all primary location with HR ranging from 1.23 to 1.55 and for metronidazole-based ATB in locally advanced rectum/junction cancer (HR=1.20 [1.04-1.40]; P=0.01).

## DISCUSSION

The present study is the first nation-wide epidemiologic cohort of patients with resected CRC where the impact of ATBs intake on recurrence has been investigated. To evaluate the link between anti-cancer drug efficacy and ATB consumption, previous studies looked at an ATBs intake 1 to 3 months before anti-cancer drug administration^15,16^. In our study, an extensive perioperative period with a range of 6 months before to 1 year after surgery was considered with the rational that ATBs can disturb gut microbiota for more than several months^17^.

Firstly, our study showed that patients who received ATBs for infections needing hospitalization after surgery were at high risk of recurrence. This finding is consistent with previous literature showing that postoperative infections are independent poor prognostic factors in CRC.^18,19^ Overall, 77% of infections were post-surgical or digestive In our study, we found an association between ATBs hospital intake and negative prognostic factors, and an excess of death during hospitalization. Therefore, the decreased 3-year DFS associated with ATBs intake during hospitalization is likely a marker of the patient’s fragility. The analysis of the impact of ATBs intake during hospitalization was also limited by the lack of access to ATB classes in this setting. Therefore, in subsequent analyses we added an additional level of control with a sensitivity analysis removing this population in order to explore the impact of the outpatient ATBs intake.

Secondly, analyses pointed out a detrimental effect of the outpatient ATBs intake after resection, mainly in locally advanced stage and when taken during chemotherapy. The increased risk of recurrence or death for post-resection ATB in this setting ranged from 22% to 41%. Antibiotics may have a differential impact depending on stage as the microbiota changes while CRC is progressing or after resection.^20^ Concerning the ATBs categories associated with an increased risk of recurrence or death, the main classes that were incriminated were penicillin A + beta-lactamase inhibitors, C3G, streptogramins, quinolones, and antifungals. Penicillin intake has been identified by others as an independent risk factor of developing CRC.^21^ Our results are in line with this evidence. C3G and quinolones have not been identified previously to increase the risk of CRC. Recent studies have identified specific fungal microbiota dysbiosis in early CRC patients compared to healthy volunteer or adenomas.^22^ Besides, fungal infections mainly occur in immunocompromised patients^23^ and underlying immunosuppression may play direct pro-tumorigenic role in CRC. Moreover, the death rate of patients receiving ATBs as outpatients was not different from patients not receiving ATBs, strengthening the idea that the observed statistical associations of ATB intake and 3-years DFS are related to the modulation of recurrence risk rather than an excess of death due to intercurrent infections.

The limitations of our study are linked to the use of a large medico-administrative database. Firstly, tumor complete pathological staging was not available to better adjust 3-year DFS. To limit biases, we reconstructed the stage based on therapeutic strategy, and performed separate analyses. Moreover, we incorporated preoperative treatments for high-risk patients and many prognostic factors into the analysis to further overcome this issue. Besides, the 6 months FOLFOX regimen was the only standard of care in the adjuvant setting in France before 2015 making the population receiving adjuvant chemotherapy homogenous. Secondly, we reconstituted the recurrence event using strong substitution parameters to obtain the outcome and its date (i.e., metastasis, palliative care, subsequent cancer surgery after resection of the primary, chemotherapy, or radiotherapy more than 3 months after resection). Consistently with other cohorts of non-metastatic CRC patients,^2^ the recurrence rate was 31% after a 3-year follow-up, supporting the substitution criteria selection in this study. Furthermore, the recurrence rate was twice as high in patients with locally advanced CRC compared to patients with localized CRC. Finally, there was a lack of consideration for prophylactic ATB during surgery. Nevertheless, all CRC patients who undergo surgery in France receive short course prophylactic ATB in a standardized way by following the national surgical guidelines^24^ and should not therefore represent a confounding factor.

Overall, this study provides evidence that ATBs could have a meaningful impact on cancer outcomes beyond the one described with immunotherapy. This observation should lead to careful indications for ATBs, limiting their number and duration in this setting as our findings indicate an association with ATBs intake and a higher risk of recurrence. Clinical studies evaluating perioperative ATBs matched with microbiota assessment in resected CRC are needed to confirm this impact and biological causality. Therapeutic strategies aiming at targeting the microbiota (e.g., dietary intervention, probiotics, fecal microbiota transplants) to reduce the CRC incidence and mortality are currently being explored.^25^

## Data Availability

The data are not publicly available due to privacy or ethical restrictions.

## Writing assistance

Michelle F.Lamendola-Essel, Magdalena Benetkiewicz (GERCOR), FRENCH, FFCD and AGEO associations are acknowledged for their manuscript reviewing.

## Author contributions

Conceptualization (MH, BR, IK, CLB, PJB, AT), Methodology (MH, BR, IK, CLB, PJB, AT, AA, CB, NG), Data validation (MH, BR, IK, CLB, PJB, AT, JV), Formal analysis (IK, CLB, PJB), Resources (IK, CLB, PJB), Data curation (IK), Writing—original draft preparation (MH), writing—review and editing (MH, BR, IK, CLB, PJB, AT, JV), Visualization (IK, BR, MH), Supervision (BR, CLB, PJB). All authors have read and agreed to the published version of the manuscript.

## Data Sharing Statement

The data are not publicly available due to privacy or ethical restrictions.

