## Supplementary Tables and Figures for "Association between the antibiotics use and recurrence in patients with resected non-metastatic colorectal cancer: EVADER-1, a nation-wide pharmaco-epidemiologic study"

### Supplementary Table 1: Classification of antibiotics.

| Antibiotic classes | ATC code | Name |
|---|---|---|
| **Azole antifungals** | J02AC01 | fluconazole |
| | J02AC02 | itraconazole |
| **Other antifungals** | J02AA01 | amphotericin B |
| | J02AX01 | flucytosine |
| **Macrolides** | J01FA01 | erythromycin |
| | J01FA02 | spiramycin |
| | J01FA03 | midecamycin |
| | J01FA06 | roxithromycin |
| | J01FA07 | josamycin |
| | J01FA09 | clarithromycin |
| | J01FA10 | azithromycin |
| | J01FA15 | telithromycin |
| **Metronidazole based-antibiotherapies** | J01RA04 | spiramycin et metronidazole |
| | P01AB01 | metronidazole |
| **Quinolones** | J01MA01 | ofloxacin |
| | J01MA02 | ciprofloxacin |
| | J01MA03 | pefloxacin |
| | J01MA04 | enoxacin |
| | J01MA06 | norfloxacin |
| | J01MA07 | lomefloxacin |
| | J01MA12 | levofloxacin |
| | J01MA14 | moxifloxacin |
| | J01MB04 | pipemidic acid |
| | J01MB07 | flumequine |
| **Streptogramins** | J01FG01 | pristinamycin |
| **Penicillin A** | J01CA04 | amoxicillin |
| **Penicillin A and beta-lactamase inhibitors** | J01CR01 | ampicillin and beta-lactamase inhibitor |
| | J01CR02 | amoxicillin and beta-lactamase inhibitor |
| **Cephalosporins (third generation)** | J01DD02 | ceftazidime |
| | J01DD04 | ceftriaxone |
| | J01DD08 | cefixime |
| | J01DD13 | cefpodoxime |
| | J01DE01 | cefepime |
| **Other beta-lactamines** | J01CA08 | pivmecillinam |
| | J01CE02 | phenoxymethylpenicillin |
| | J01CE08 | benzathine benzylpenicillin |
| | J01CF02 | cloxacillin |
| | J01CF04 | oxacillin |

| | | |
|---|---|---|
| | J01CR03 | ticarcillin and beta-lactamase inhibitor |
| | J01CR05 | piperacillin and beta-lactamase inhibitor |
| | J01DB01 | cefalexin |
| | J01DB05 | cefadroxil |
| | J01DC02 | cefuroxime |
| | J01DC04 | cefaclor |
| | J01DC07 | cefotiam |
| | J01DF01 | aztreonam |
| | J01DH02 | meropenem |
| | J01DH51 | imipenem and cilastatin |
| **Other antibiotics** | J01AA02 | doxycycline |
| | J01AA05 | metacycline |
| | J01AA07 | tetracycline |
| | J01AA08 | minocycline |
| | J01BA02 | thiamphenicol |
| | J01EE01 | sulfamethoxazole and trimethoprim |
| | J01FF01 | clindamycin |
| | J01FF02 | lincomycin |
| | J01GB01 | tobramycin |
| | J01GB07 | netilmicin |
| | J01XB01 | colistin |
| | J01XC01 | fusidic acid |
| | J01XE01 | nitrofurantoin |
| | J01XX01 | fosfomycin |
| | J04AB02 | rifampicin |
| | J04AB04 | rifabutin |
| | J04AC01 | isoniazid |
| | J04AK01 | pyrazinamide |
| | J04AK02 | ethambutol |
| | J04AM02 | rifampicin and isoniazid |

**Supplementary Table 2. Description of most frequently prescribed antibiotic combinations (other than other) in patients who received several antibiotics (n=13 669).**

| ATB Combinations | n | % |
|---|---|---|
| Penicillin A and beta-lactamase inhibitors + Quinolones | 625 | 4.6 |
| Penicillin A + Penicillin A and beta-lactamase inhibitors | 505 | 3.7 |
| Metronidazole based-antibiotherapies + Quinolones | 450 | 3.3 |
| Penicillin A + Quinolones | 449 | 3.3 |
| Penicillin A and beta-lactamase inhibitors + Metronidazole based-antibiotherapies | 376 | 2.8 |
| Cephalosporins (third generation) + Quinolones | 344 | 2.6 |
| Penicillin A + Macrolides | 321 | 2.4 |
| Cephalosporins (third generation) + Penicillin A and beta-lactamase inhibitors + Quinolones | 289 | 2.2 |
| Cephalosporins (third generation) + Penicillin A Quinolones | 282 | 2.1 |
| Penicillin A + Macrolides + Metronidazole based-antibiotherapies | 260 | 1.9 |
| Other combinations | 9 768 | 71.5 |

**Supplementary Table 3. Infections requiring antibiotics at hospital during the perioperative period (n= 13 319 patients).**

| | Within 6 months before surgery | During surgical resection stay | 1-30 days after surgery | 31-90 days after surgery | 91 days-1 year after surgery | Total | |
|---|---|---|---|---|---|---|---|
| | n | n | n | n | n | n | % |
| **Bacteremias** | 158 | 854 | 116 | 126 | 316 | 1 471 | 11.0 |
| **Digestive infections** | 1 429 | 3 368 | 555 | 406 | 632 | 5 574 | 41.8 |
| **Post-surgical site infections** | 132 | 3 800 | 672 | 563 | 642 | 5 125 | 38.5 |
| **Respiratory infections** | 310 | 1 190 | 188 | 206 | 542 | 2 201 | 16.5 |
| **Urinary infections** | 81 | 170 | 48 | 101 | 135 | 504 | 3.8 |
| **Bone infections** | 38 | 9 | 4 | 18 | 29 | 86 | 0.6 |
| **Cutaneous infections** | 65 | 105 | 18 | 20 | 70 | 265 | 2.0 |
| **Not specified infections** | 345 | 1 979 | 124 | 155 | 257 | 2 270 | 17.0 |
| **Total *** | 2 576 | 9 540 | 1 294 | 1 377 | 2 300 | 13 326 | 100 |

*One patient may have multiple infections thus the total is not necessarily equal to the sum of all patients.

**Supplementary table 4. Pre- and post resection exposure to antibiotics per class in the cohort of patients with resected colorectal cancer.**

| | Period | Number of patients | Number of days | % of days of the period | Nb of patients with microbiota impact during chemo | Number of days with microbiota impact during chemo | % of days with microbiota impact during chemo |
|---|---|---|---|---|---|---|---|
| | | n | median (q1-q3) | median (q1-q3) | n | median (q1-q3) | median (q1-q3) |
| Global | Pre resection | 13 779 | 43 (34-61) | 24 (19-34) | 1 261 | 28 (16-40) | 61 (34-92) |
| Global | Post resection | 21 601 | 55 (39-95) | 18 (11-30) | 5 112 | 45 (31-76) | 33 (20-56) |
| C3G | Pre resection | 1 755 | 36 (32-39) | 20 (18-22) | 110 | 20 (10-30) | 40.5 (23-69) |
| C3G | Post resection | 3 109 | 36 (33-41) | 10 (9-13) | 494 | 35 (24-39) | 21 (15-26) |
| Antifungal – Azoles | Pre resection | 459 | 34 (31-38) | 19 (17-21) | 103 | 26 (17-33) | 51 (31-74) |
| Antifungal – Azoles | Post resection | 1 531 | 34 (32-50) | 10 (9-18) | 804 | 34 (30.5-57.5) | 23 (18-42) |
| Antifungal – Others | Pre resection | 559 | 38 (33-47) | 21 (18-26) | 195 | 32 (25-42) | 64 (37-93) |
| Antifungal – Others | Post resection | 2 252 | 40 (34-64) | 12 (10-21) | 1 436 | 39 (32-69) | 29 (20-52) |
| Beta-lactamines – Others | Pre resection | 691 | 38 (33-42) | 21 (18-23) | 33 | 21 (15-29) | 57 (33-81) |
| Beta-lactamines – Others | Post resection | 1 445 | 39 (35-45) | 11 (10-14) | 250 | 38 (27-46) | 23 (16-36) |
| Penicillin A | Pre resection | 3 815 | 43 (35-49) | 24 (19-28) | 260 | 25 (15-36) | 54 (28-84) |
| Penicillin A | Post resection | 6 556 | 45 (39-55) | 12 (11-16) | 1 031 | 43 (31-49) | 26 (19-33) |
| Penicillin A + beta-lactamase inhibitor | Pre resection | 3 229 | 46 (31-55) | 26 (17-31) | 301 | 30 (16-41) | 55 (34-89) |
| Penicillin A + beta-lactamase inhibitor | Post resection | 7 132 | 52 (42-58) | 15 (12-20) | 1 649 | 42 (22-54) | 27 (16-35) |
| Macrolides | Pre resection | 2 171 | 36 (31-41) | 20 (17-23) | 120 | 24 (15-31.5) | 53 (28.5-81.5) |
| Macrolides | Post resection | 3 015 | 36 (33-41) | 10 (9-12) | 358 | 36 (23-41) | 21 (14-26) |
| Metronidazole based-antibiotherapies | Pre resection | 2 933 | 33 (21-37) | 18 (12-21) | 161 | 21 (12-30) | 41 (25-65) |

| | | | | | | | |
|---|---|---|---|---|---|---|---|
| Metronidazole based-antibiotherapies | Post resection | 3 910 | 34 (29-38) | 9 (8-11) | 440 | 27 (12-34) | 17 (9-21) |
| Quinolones | Pre resection | 3 286 | 37 (33-43) | 21 (19-24) | 347 | 25 (13-36) | 43 (25-69) |
| Quinolones | Post resection | 7 342 | 40 (36-46) | 11 (10-17) | 1 579 | 35 (18-41) | 22 (13-30) |
| Streptogramins | Pre resection | 858 | 39 (35-42) | 22 (19-24) | 47 | 26 (11-38) | 45 (23-73) |
| Streptogramins | Post resection | 1 606 | 39 (36-43) | 11 (10-13) | 251 | 37 (23-42) | 22 (14-27) |
| Other ATB including cyclins | Pre resection | 1 707 | 36 (31-52) | 20 (17-29) | 168 | 32.5 (12.5-59.5) | 51.5 (26-84) |
| Other ATB including cyclins | Post resection | 3 387 | 38 (32-59) | 11 (9-19) | 591 | 36 (22-61) | 24 (16-45) |
| Infections needing hospitalization | Pre resection | 2 557 | 7 (1-14) | 4 (1-8) | 60 | 10.5 (6-16.5) | 17.5 (8.5-44) |
| Infections needing hospitalization | Post resection | 11 901 | 18 (12-30) | 6 (4-13) | 455 | 8 (3-15) | 7 (3-32) |

**Supplementary table 5. Days of adjuvant chemotherapy with antibiotic intake or impact on the microbiota.**

How to read the table: 409 patients (4.4% of patients with adjuvant chemotherapy) had between 1 and 9% of their chemotherapy period time with an impact on microbiota related to any ATB.

| Days | ATB as outpatient (all ATB) | | C3G | | Antifungal - Other | | Antifungal Azoles | | Other betalactamine s | | Penicillin A | | Penicillin A + beta-lactamase inhibitors | | Macrolides | | Metronidazol e based-ATB | | Quinolones | | Streptogramins | | Other ATB including cyclins | |
|---|---|---|---|---|---|---|---|---|---|---|---|---|---|---|---|---|---|---|---|---|---|---|---|---|
| | n | % | n | % | n | % | n | % | n | % | n | % | n | % | n | % | n | % | n | % | n | % | n | % |
| **0** | 4 123 | 44.7 | 8 738 | 94.6 | 7 797 | 84.5 | 8 428 | 91.3 | 8 982 | 97.3 | 8 203 | 88.9 | 7 584 | 82.1 | 8 875 | 96.1 | 8 795 | 95.3 | 7 655 | 82.9 | 8 982 | 97.3 | 8 641 | 93.6 |
| **01-09** | 409 | 4.4 | 67 | 0.7 | 70 | 0.8 | 44 | 0.5 | 31 | 0.3 | 111 | 1.2 | 213 | 2.3 | 48 | 0.5 | 115 | 1.2 | 265 | 2.9 | 32 | 0.3 | 84 | 0.9 |
| **10-19** | 731 | 7.9 | 144 | 1.6 | 254 | 2.8 | 223 | 2.4 | 62 | 0.7 | 153 | 1.7 | 328 | 3.6 | 94 | 1.0 | 153 | 1.7 | 389 | 4.2 | 67 | 0.7 | 128 | 1.4 |
| **20-29** | 1 085 | 11.8 | 176 | 1.9 | 400 | 4.3 | 231 | 2.5 | 83 | 0.9 | 393 | 4.3 | 432 | 4.7 | 161 | 1.7 | 107 | 1.2 | 517 | 5.6 | 98 | 1.1 | 134 | 1.5 |
| **30-39** | 789 | 8.5 | 39 | 0.4 | 189 | 2.0 | 89 | 1.0 | 16 | 0.2 | 209 | 2.3 | 337 | 3.7 | 26 | 0.3 | 27 | 0.3 | 136 | 1.5 | 20 | 0.2 | 70 | 0.8 |
| **40-49** | 547 | 5.9 | 32 | 0.3 | 144 | 1.6 | 56 | 0.6 | 9 | 0.1 | 67 | 0.7 | 112 | 1.2 | 8 | 0.1 | 19 | 0.2 | 102 | 1.1 | 14 | 0.2 | 39 | 0.4 |
| **50-59** | 392 | 4.2 | 13 | 0.1 | 89 | 1.0 | 48 | 0.5 | 4 | 0.0 | 41 | 0.4 | 60 | 0.6 | 4 | 0.0 | 3 | 0.0 | 53 | 0.6 | 5 | 0.1 | 31 | 0.3 |
| **60-69** | 310 | 3.4 | 7 | 0.1 | 69 | 0.7 | 26 | 0.3 | 2 | 0.0 | 25 | 0.3 | 50 | 0.5 | 6 | 0.1 | 4 | 0.0 | 30 | 0.3 | 4 | 0.0 | 19 | 0.2 |
| **70-79** | 218 | 2.4 | 7 | 0.1 | 54 | 0.6 | 15 | 0.2 | 3 | 0.0 | 4 | 0.0 | 26 | 0.3 | 1 | 0.0 | 2 | 0.0 | 19 | 0.2 | 3 | 0.0 | 18 | 0.2 |
| **80-89** | 164 | 1.8 | 2 | 0.0 | 39 | 0.4 | 14 | 0.2 | 7 | 0.1 | 8 | 0.1 | 20 | 0.2 | 3 | 0.0 | 2 | 0.0 | 10 | 0.1 | 1 | 0.0 | 20 | 0.2 |
| **90-100** | 464 | 5.0 | 7 | 0.1 | 127 | 1.4 | 58 | 0.6 | 33 | 0.4 | 18 | 0.2 | 70 | 0.8 | 6 | 0.1 | 5 | 0.1 | 56 | 0.6 | 6 | 0.1 | 48 | 0.5 |

### Supplementary Table 6. Multivariate analyses for 3-year DFS in patients with resected colorectal cancer according to antibiotic and chemotherapy exposure.

#### 6A-Localized colon cancer patients (n=17,335)

| Analysis of Maximum Likelihood Estimates | | | | | |
|---|---|---|---|---|---|
| **Parameter** | | **Pr > ChiSq** | **Hazard Ratio** | **95% Hazard Ratio Confidence Limits** | |
| Sex | Male *vs* Female | <.0001 | 1.152 | 1.092 | 1.215 |
| Age | 50 to 69 *vs* <50 | 0.0950 | 1.146 | 0.977 | 1.344 |
| | 70-79 *vs* <50 | <.0001 | 1.444 | 1.230 | 1.695 |
| | ≥ 80 *vs* <50 | <.0001 | 2.276 | 1.946 | 2.663 |
| Charlson | 1 à 2 *vs* 0 | <.0001 | 1.144 | 1.077 | 1.215 |
| | ≥ 3 *vs* 0 | <.0001 | 1.569 | 1.459 | 1.688 |
| Malnutrition | Yes *vs* No | <.0001 | 1.363 | 1.273 | 1.459 |
| ICU stay | > 2 to 7 days *vs* 0-2 days | <.0001 | 1.485 | 1.330 | 1.657 |
| | > 7 days *vs* 0-2 days | <.0001 | 2.196 | 1.987 | 2.428 |
| *Pre resection ATB exposure* | | | | | |
| ATB as outpatient | Yes *vs* No | 0.9665 | 0.999 | 0.944 | 1.057 |
| ATB with hospitalization | Yes *vs* No | 0.8671 | 1.008 | 0.920 | 1.104 |
| *Post resection ATB exposure* | | | | | |
| ATB as outpatient | Yes *vs* No | 0.1471 | 1.045 | 0.985 | 1.108 |
| **ATB with hospitalization ≤ 30 days after resection** | **Yes *vs* No** | **<.0001** | **1.614** | **1.524** | **1.709** |
| **ATB with hospitalization > 30 days after resection** | **Yes *vs* No** | **<.0001** | **2.341** | **2.058** | **2.663** |

| Analysis of Maximum Likelihood Estimates | | | | | |
|---|---|---|---|---|---|
| **Parameter** | | **Pr > ChiSq** | **Hazard Ratio** | **95% Hazard Ratio Confidence Limits** | |
| Sex | Male *vs* Female | <.0001 | 1.151 | 1.091 | 1.214 |
| Age | 50 to 69 *vs* <50 | 0.1026 | 1.142 | 0.974 | 1.341 |
| | 70-79 *vs* <50 | <.0001 | 1.443 | 1.229 | 1.694 |
| | ≥ 80 *vs* <50 | <.0001 | 2.272 | 1.941 | 2.658 |
| Charlson | 1 à 2 *vs* 0 | <.0001 | 1.141 | 1.074 | 1.212 |
| | ≥ 3 *vs* 0 | <.0001 | 1.559 | 1.450 | 1.678 |
| Malnutrition | Yes *vs* No | <.0001 | 1.361 | 1.272 | 1.457 |
| ICU stay | > 2 to 7 days *vs* 0-2 days | <.0001 | 1.483 | 1.328 | 1.656 |
| | > 7 days *vs* 0-2 days | <.0001 | 2.208 | 1.997 | 2.441 |
| *Pre resection ATB exposure* | | | | | |
| ATB as outpatient | Yes *vs* No | 0.6928 | 0.989 | 0.934 | 1.046 |
| ATB with hospitalization | Yes *vs* No | 0.7395 | 1.016 | 0.927 | 1.113 |
| *Post resection ATB exposure* | | | | | |
| C3G | Yes *vs* No | 0.5794 | 1.028 | 0.932 | 1.135 |
| **Antifungal – Others** | Yes *vs* No | **0.0010** | **1.311** | **1.115** | **1.542** |
| Antifungal – Azoles | Yes *vs* No | 0.0468 | 1.190 | 1.002 | 1.412 |
| Beta-lactamines – Others | Yes *vs* No | 0.7243 | 1.026 | 0.890 | 1.183 |
| Penicillin A | Yes *vs* No | 0.4513 | 0.970 | 0.896 | 1.050 |
| **Penicillin A + beta-lactamase inhibitor** | Yes *vs* No | **0.0097** | **1.102** | **1.024** | **1.186** |

| | | | | | |
|---|---|---|---|---|---|
| **Macrolides** | Yes *vs* No | **0.0344** | **0.887** | **0.794** | **0.991** |
| Metronidazole based-antibiotherapies | Yes *vs* No | 0.6903 | 1.021 | 0.923 | 1.128 |
| Quinolones | Yes *vs* No | 0.2057 | 1.049 | 0.974 | 1.129 |
| **Streptogramins** | Yes *vs* No | **0.0070** | **1.197** | **1.050** | **1.365** |
| Other ATB including cyclins | Yes *vs* No | 0.5505 | 1.031 | 0.933 | 1.140 |
| **ATB with hospitalization ≤ 30 days after resection** | Yes *vs* No | **<.0001** | **1.590** | **1.501** | **1.685** |
| **ATB with hospitalization > 30 days after resection** | Yes *vs* No | **<.0001** | **2.266** | **1.990** | **2.581** |

### 6B-Advanced colon cancer patients (n=5,805)

| Analysis of Maximum Likelihood Estimates | | | | | |
|---|---|---|---|---|---|
| **Parameter** | | **Pr > ChiSq** | **Hazard Ratio** | **95% Hazard Ratio Confidence Limits** | |
| Sex | Male *vs* Female | <.0001 | 1.227 | 1.129 | 1.333 |
| Age | 50 to 69 *vs* <50 | 0.8100 | 1.019 | 0.877 | 1.183 |
| | 70-79 *vs* <50 | 0.3136 | 1.085 | 0.926 | 1.273 |
| | ≥ 80 *vs* <50 | <.0001 | 1.478 | 1.235 | 1.769 |
| Charlson | 1 à 2 *vs* 0 | 0.3566 | 1.047 | 0.950 | 1.153 |
| | ≥ 3 *vs* 0 | 0.0601 | 1.171 | 0.993 | 1.381 |
| Malnutrition | Yes *vs* No | 0.0057 | 1.183 | 1.050 | 1.333 |
| ICU stay | > 2 to 7 days *vs* 0-2 days | 0.1068 | 1.178 | 0.965 | 1.439 |
| | > 7 days *vs* 0-2 days | <.0001 | 2.000 | 1.601 | 2.498 |
| *Pre resection ATB exposure* | | | | | |
| ATB as outpatient | Yes *vs* No | 0.5915 | 1.024 | 0.938 | 1.118 |
| ATB with hospitalization | Yes *vs* No | 0.7785 | 1.022 | 0.879 | 1.189 |
| *Post resection ATB exposure* | | | | | |
| **ATB as outpatient** | **Yes *vs* No** | **0.0001** | **1.201** | **1.093** | **1.321** |
| **ATB with hospitalization ≤ 30 days after resection** | **Yes *vs* No** | **<.0001** | **1.305** | **1.192** | **1.428** |
| **ATB with hospitalization > 30 days after resection** | **Yes *vs* No** | **<.0001** | **1.675** | **1.395** | **2.011** |

| Analysis of Maximum Likelihood Estimates | | | | | |
|---|---|---|---|---|---|
| **Parameter** | | **Pr > ChiSq** | **Hazard Ratio** | **95% Hazard Ratio Confidence Limits** | |
| Sex | Male *vs* Female | <.0001 | 1.210 | 1.113 | 1.315 |
| Age | 50 to 69 *vs* <50 | 0.7850 | 1.021 | 0.879 | 1.186 |
| | 70-79 *vs* <50 | 0.2585 | 1.096 | 0.935 | 1.286 |
| | ≥ 80 *vs* <50 | <.0001 | 1.500 | 1.251 | 1.797 |
| Charlson | 1 à 2 *vs* 0 | 0.3216 | 1.050 | 0.953 | 1.157 |
| | ≥ 3 *vs* 0 | 0.2074 | 1.113 | 0.942 | 1.314 |
| Malnutrition | Yes *vs* No | 0.0043 | 1.190 | 1.056 | 1.340 |
| ICU stay | > 2 to 7 days *vs* 0-2 days | 0.1331 | 1.165 | 0.954 | 1.423 |
| | > 7 days *vs* 0-2 days | <.0001 | 1.864 | 1.491 | 2.332 |
| *Pre resection ATB exposure* | | | | | |
| ATB as outpatient | ATB/no neoadjuvant *vs* no ATB/no neoadjuvant | 0.4730 | 0.967 | 0.882 | 1.060 |
| Timing of chemotherapy and antibiotherapy | ATB outside neoadjuvant *vs* Neoadjuvant/no ATB | 0.5350 | 1.204 | 0.670 | 2.164 |

| | | | | | |
|---|---|---|---|---|---|
| | ATB during neoadjuvant *vs* Neoadjuvant/no ATB | 0.0706 | 1.353 | 0.975 | 1.878 |
| ATB with hospitalization | Yes *vs* No | 0.8349 | 0.984 | 0.846 | 1.145 |
| *Post resection ATB exposure* | | | | | |
| **ATB outside chemotherapy** | **Yes *vs* No** | **0.0256** | **1.164** | **1.019** | **1.330** |
| **ATB during chemotherapy** | **Yes *vs* No** | **<.0001** | **1.224** | **1.110** | **1.350** |
| **ATB with hospitalization ≤ 30 days after resection** | **Yes *vs* No** | **<.0001** | **1.300** | **1.188** | **1.423** |
| **ATB with hospitalization > 30 days after resection** | **Yes *vs* No** | **<.0001** | **1.710** | **1.423** | **2.053** |

| Analysis of Maximum Likelihood Estimates | | | | | |
|---|---|---|---|---|---|
| **Parameter** | | **Pr > ChiSq** | **Hazard Ratio** | **95% Hazard Ratio Confidence Limits** | |
| Sex | Male *vs* Female | <.0001 | 1.246 | 1.146 | 1.355 |
| Age | 50 to 69 *vs* <50 | 0.7386 | 1.026 | 0.883 | 1.192 |
| | 70-79 *vs* <50 | 0.2924 | 1.090 | 0.929 | 1.278 |
| | ≥ 80 *vs* <50 | <.0001 | 1.493 | 1.246 | 1.789 |
| Charlson | 1 à 2 *vs* 0 | 0.4734 | 1.036 | 0.940 | 1.142 |
| | ≥ 3 *vs* 0 | 0.2064 | 1.113 | 0.942 | 1.315 |
| Malnutrition | Yes *vs* No | 0.0032 | 1.197 | 1.062 | 1.349 |
| ICU stay | > 2 to 7 days *vs* 0-2 days | 0.1384 | 1.164 | 0.952 | 1.422 |
| | > 7 days *vs* 0-2 days | <.0001 | 1.840 | 1.470 | 2.302 |
| *Pre resection ATB exposure* | | | | | |
| ATB as outpatient | ATB/no neoadjuvant *vs* no ATB/no neoadjuvant | 0.2836 | 0.950 | 0.866 | 1.043 |
| Timing of chemotherapy and antibiotherapy | ATB outside neoadjuvant *vs* Neoadjuvant/no ATB | 0.5895 | 1.176 | 0.653 | 2.117 |
| | ATB during neoadjuvant *vs* Neoadjuvant/no ATB | 0.1936 | 1.244 | 0.895 | 1.728 |
| ATB with hospitalization | Yes *vs* No | 0.9269 | 0.993 | 0.853 | 1.156 |
| *Post resection ATB exposure* | | | | | |
| **C3G** | Yes *vs* No | **0.0099** | **1.203** | **1.045** | **1.385** |
| **Antifungal – Others** | Yes *vs* No | **0.0289** | **1.126** | **1.012** | **1.253** |
| Antifungal – Azoles | Yes *vs* No | 0.5483 | 1.043 | 0.910 | 1.194 |
| Beta-lactamines – Others | Yes *vs* No | 0.1369 | 0.855 | 0.696 | 1.051 |
| Penicillin A | Yes *vs* No | 0.1637 | 1.079 | 0.969 | 1.201 |
| Penicillin A + beta-lactamase inhibitor | Yes *vs* No | 0.6340 | 1.024 | 0.930 | 1.127 |
| Macrolides | Yes *vs* No | 0.4734 | 0.944 | 0.806 | 1.105 |
| Metronidazole based-antibiotherapies | Yes *vs* No | 0.0590 | 0.877 | 0.766 | 1.005 |
| **Quinolones** | Yes *vs* No | **0.0093** | **1.139** | **1.033** | **1.257** |
| **Streptogramins** | Yes *vs* No | **0.0240** | **1.229** | **1.027** | **1.470** |
| **Other ATB including cyclins** | Yes *vs* No | **<.0001** | **1.490** | **1.310** | **1.695** |
| **ATB with hospitalization ≤ 30 days after resection** | Yes *vs* No | **<.0001** | **1.283** | **1.171** | **1.405** |
| **ATB with hospitalization > 30 days after resection** | Yes *vs* No | **<.0001** | **1.639** | **1.361** | **1.972** |

### 6C-Localized rectum/junction cancer (N=5,097)

| Analysis of Maximum Likelihood Estimates | | | | | |
|---|---|---|---|---|---|
| **Parameter** | | **Pr > ChiSq** | **Hazard Ratio** | **95% Hazard Ratio Confidence Limits** | |
| Sex | Male *vs* Female | <.0001 | 1.338 | 1.214 | 1.475 |
| Age | 50 to 69 *vs* <50 | 0.6873 | 1.056 | 0.809 | 1.379 |
| | 70-79 *vs* <50 | 0.0292 | 1.354 | 1.031 | 1.777 |
| | ≥ 80 *vs* <50 | <.0001 | 1.877 | 1.434 | 2.458 |
| Charlson | 1 à 2 *vs* 0 | 0.0287 | 1.130 | 1.013 | 1.260 |
| | ≥ 3 *vs* 0 | <.0001 | 1.448 | 1.249 | 1.678 |
| Malnutrition | Yes *vs* No | 0.0025 | 1.249 | 1.081 | 1.444 |
| ICU stay | > 2 to 7 days *vs* 0-2 days | 0.0090 | 1.301 | 1.068 | 1.585 |
| | > 7 days *vs* 0-2 days | <.0001 | 1.880 | 1.543 | 2.291 |
| *Pre resection ATB exposure* | | | | | |
| ATB as outpatient | Yes *vs* No | 0.6897 | 0.979 | 0.883 | 1.086 |
| ATB with hospitalization | Yes *vs* No | 0.7470 | 1.032 | 0.854 | 1.246 |
| *Post resection ATB exposure* | | | | | |
| ATB as outpatient | Yes *vs* No | 0.4120 | 0.957 | 0.861 | 1.063 |
| **ATB with hospitalization ≤ 30 days after resection** | **Yes *vs* No** | **<.0001** | **1.845** | **1.664** | **2.047** |
| **ATB with hospitalization > 30 days after resection** | **Yes *vs* No** | **<.0001** | **2.437** | **1.962** | **3.027** |

| Analysis of Maximum Likelihood Estimates | | | | | |
|---|---|---|---|---|---|
| **Parameter** | | **Pr > ChiSq** | **Hazard Ratio** | **95% Hazard Ratio Confidence Limits** | |
| Sex | Male *vs* Female | <.0001 | 1.328 | 1.204 | 1.464 |
| Age | 50 to 69 *vs* <50 | 0.6336 | 1.067 | 0.817 | 1.393 |
| | 70-79 *vs* <50 | 0.0211 | 1.378 | 1.049 | 1.809 |
| | ≥ 80 *vs* <50 | <.0001 | 1.909 | 1.457 | 2.500 |
| Charlson | 1 à 2 *vs* 0 | 0.0440 | 1.119 | 1.003 | 1.249 |
| | ≥ 3 *vs* 0 | <.0001 | 1.443 | 1.243 | 1.674 |
| Malnutrition | Yes *vs* No | 0.0020 | 1.256 | 1.087 | 1.451 |
| ICU stay | > 2 to 7 days *vs* 0-2 days | 0.0075 | 1.309 | 1.075 | 1.595 |
| | > 7 days *vs* 0-2 days | <.0001 | 1.935 | 1.586 | 2.360 |
| *Pre resection ATB exposure* | | | | | |
| ATB as outpatient | Yes *vs* No | 0.3461 | 0.951 | 0.858 | 1.055 |
| ATB with hospitalization | Yes *vs* No | 0.5880 | 1.054 | 0.872 | 1.274 |
| *Post resection ATB exposure* | | | | | |
| C3G | Yes *vs* No | 0.1946 | 1.136 | 0.937 | 1.378 |
| Antifungal – Others | Yes *vs* No | 0.6362 | 1.079 | 0.787 | 1.480 |
| Antifungal – Azoles | Yes *vs* No | 0.2869 | 1.208 | 0.853 | 1.711 |
| Beta-lactamines – Others | Yes *vs* No | 0.5748 | 0.919 | 0.684 | 1.235 |
| Penicillin A | Yes *vs* No | 0.6333 | 0.966 | 0.837 | 1.114 |
| **Penicillin A + beta-lactamase inhibitor** | Yes *vs* No | **0.0070** | **1.200** | **1.051** | **1.370** |

| | | | | | |
|---|---|---|---|---|---|
| **Macrolides** | Yes *vs* No | **0.0267** | **0.788** | **0.638** | **0.973** |
| Metronidazole based-antibiotherapies | Yes *vs* No | 0.8949 | 0.989 | 0.839 | 1.166 |
| Quinolones | Yes *vs* No | 0.9377 | 1.005 | 0.882 | 1.146 |
| Streptogramins | Yes *vs* No | 0.2027 | 0.843 | 0.649 | 1.096 |
| Other ATB including cyclins | Yes *vs* No | 0.4060 | 1.077 | 0.904 | 1.284 |
| **ATB with hospitalization ≤ 30 days after resection** | Yes *vs* No | **<.0001** | **1.784** | **1.605** | **1.983** |
| **ATB with hospitalization > 30 days after resection** | Yes *vs* No | **<.0001** | **2.337** | **1.877** | **2.910** |

### 6D-Advanced rectum/junction cancer patients (N=6,859)

**Analysis of Maximum Likelihood Estimates**

| **Parameter** | | **Pr > ChiSq** | **Hazard Ratio** | **95% Hazard Ratio Confidence Limits** | |
|---|---|---|---|---|---|
| Sex | Male *vs* Female | <.0001 | 1.148 | 1.072 | 1.229 |
| Age | 50 to 69 *vs* <50 | 0.4649 | 0.957 | 0.850 | 1.077 |
| | 70-79 *vs* <50 | 0.7972 | 0.983 | 0.864 | 1.119 |
| | ≥ 80 *vs* <50 | 0.0941 | 1.132 | 0.979 | 1.309 |
| Charlson | 1 à 2 *vs* 0 | 0.0959 | 1.068 | 0.988 | 1.153 |
| | ≥ 3 *vs* 0 | 0.3683 | 1.068 | 0.926 | 1.231 |
| Malnutrition | Yes *vs* No | 0.0009 | 1.170 | 1.067 | 1.284 |
| ICU stay | > 2 to 7 days *vs* 0-2 days | 0.3122 | 0.920 | 0.783 | 1.081 |
| | > 7 days *vs* 0-2 days | 0.0192 | 1.238 | 1.035 | 1.481 |
| *Pre resection ATB exposure* | | | | | |
| ATB as outpatient | Yes *vs* No | 0.0735 | 1.065 | 0.994 | 1.141 |
| ATB with hospitalization | Yes *vs* No | 0.8750 | 0.989 | 0.861 | 1.136 |
| *Post resection ATB exposure* | | | | | |
| ATB as outpatient | Yes *vs* No | 0.1026 | 1.062 | 0.988 | 1.142 |
| **ATB with hospitalization ≤ 30 days after resection** | **Yes *vs* No** | **<.0001** | **1.269** | **1.182** | **1.363** |
| **ATB with hospitalization > 30 days after resection** | **Yes *vs* No** | **<.0001** | **1.642** | **1.438** | **1.877** |

**Analysis of Maximum Likelihood Estimates**

| **Parameter** | | **Pr > ChiSq** | **Hazard Ratio** | **95% Hazard Ratio Confidence Limits** | |
|---|---|---|---|---|---|
| Sex | Male *vs* Female | 0.0004 | 1.133 | 1.058 | 1.213 |
| Age | 50 to 69 *vs* <50 | 0.6802 | 0.975 | 0.866 | 1.098 |
| | 70-79 *vs* <50 | 0.6609 | 1.029 | 0.904 | 1.172 |
| | ≥ 80 *vs* <50 | 0.0005 | 1.303 | 1.124 | 1.510 |
| Charlson | 1 à 2 *vs* 0 | 0.0353 | 1.086 | 1.006 | 1.174 |
| | ≥ 3 *vs* 0 | 0.1816 | 1.102 | 0.956 | 1.271 |
| Malnutrition | Yes *vs* No | 0.0021 | 1.156 | 1.054 | 1.268 |
| ICU stay | > 2 to 7 days *vs* 0-2 days | 0.2898 | 0.917 | 0.780 | 1.077 |
| | > 7 days *vs* 0-2 days | 0.0114 | 1.260 | 1.053 | 1.506 |
| *Pre resection ATB exposure* | | | | | |

| | | | | | |
|---|---|---|---|---|---|
| ATB as outpatient | ATB/no neoadjuvant *vs* no ATB/no neoadjuvant | 0.2816 | 1.071 | 0.945 | 1.214 |
| Timing of chemotherapy and antibiotherapy | ATB outside neoadjuvant *vs* Neoadjuvant/no ATB | 0.3115 | 0.943 | 0.842 | 1.056 |
| | **ATB during neoadjuvant *vs* Neoadjuvant/no ATB** | **0.0089** | **1.132** | **1.032** | **1.242** |
| ATB with hospitalization | Yes *vs* No | 0.9170 | 1.007 | 0.877 | 1.157 |
| *Post resection ATB exposure* | | | | | |
| ATB outside chemotherapy | Yes *vs* No | 0.1353 | 0.939 | 0.865 | 1.020 |
| **ATB during chemotherapy** | **Yes *vs* No** | **<.0001** | **1.312** | **1.202** | **1.432** |
| **ATB with hospitalization ≤ 30 days after resection** | **Yes *vs* No** | **<.0001** | **1.257** | **1.170** | **1.351** |
| **ATB with hospitalization > 30 days after resection** | **Yes *vs* No** | **<.0001** | **1.604** | **1.403** | **1.833** |

**Analysis of Maximum Likelihood Estimates**

| **Parameter** | | **Pr > ChiSq** | **Hazard Ratio** | **95% Hazard Ratio Confidence Limits** | |
|---|---|---|---|---|---|
| Sex | Male *vs* Female | <.0001 | 1.147 | 1.071 | 1.229 |
| Age | 50 to 69 *vs* <50 | 0.6087 | 0.969 | 0.861 | 1.092 |
| | 70-79 *vs* <50 | 0.9365 | 1.005 | 0.883 | 1.145 |
| | ≥ 80 *vs* <50 | 0.0060 | 1.229 | 1.061 | 1.423 |
| Charlson | 1 à 2 *vs* 0 | 0.0602 | 1.077 | 0.997 | 1.164 |
| | ≥ 3 *vs* 0 | 0.3099 | 1.077 | 0.934 | 1.242 |
| Malnutrition | Yes *vs* No | 0.0015 | 1.162 | 1.059 | 1.275 |
| ICU stay | > 2 to 7 days *vs* 0-2 days | 0.1941 | 0.899 | 0.765 | 1.056 |
| | > 7 days *vs* 0-2 days | 0.0199 | 1.237 | 1.034 | 1.480 |
| *Pre resection ATB exposure* | | | | | |
| ATB as outpatient | ATB/no neoadjuvant *vs* no ATB/no neoadjuvant | 0.4313 | 1.052 | 0.927 | 1.195 |
| Timing of chemotherapy and antibiotherapy | ATB outside neoadjuvant *vs* Neoadjuvant/no ATB | 0.1330 | 0.917 | 0.819 | 1.027 |
| | **ATB during neoadjuvant *vs* Neoadjuvant/no ATB** | **0.0294** | **1.109** | **1.010** | **1.218** |
| ATB with hospitalization | Yes *vs* No | 0.9935 | 1.001 | 0.871 | 1.149 |
| *Post resection ATB exposure* | | | | | |
| **C3G** | **Yes *vs* No** | **0.0008** | **1.223** | **1.088** | **1.376** |
| Antifungal – Others | Yes *vs* No | 0.1863 | 1.083 | 0.962 | 1.219 |
| **Antifungal – Azoles** | **Yes *vs* No** | **0.0021** | **1.236** | **1.080** | **1.415** |
| Beta-lactamines – Others | Yes *vs* No | 0.8044 | 1.023 | 0.852 | 1.230 |
| Penicillin A | Yes *vs* No | 0.5429 | 0.970 | 0.881 | 1.069 |
| Penicillin A + beta-lactamase inhibitor | Yes *vs* No | 0.4198 | 1.035 | 0.952 | 1.126 |
| Macrolides | Yes *vs* No | 0.6294 | 0.964 | 0.831 | 1.119 |
| Metronidazole based-antibiotherapies | Yes *vs* No | 0.0604 | 1.104 | 0.996 | 1.224 |
| Quinolones | Yes *vs* No | 0.3096 | 0.959 | 0.885 | 1.039 |
| Streptogramins | Yes *vs* No | 0.0665 | 1.166 | 0.990 | 1.375 |
| **Other ATB including cyclins** | **Yes *vs* No** | **0.0001** | **1.234** | **1.109** | **1.372** |
| **ATB with hospitalization ≤ 30 days after resection** | **Yes *vs* No** | **<.0001** | **1.232** | **1.146** | **1.325** |
| **ATB with hospitalization > 30 days after resection** | **Yes *vs* No** | **<.0001** | **1.542** | **1.347** | **1.766** |

**Supplementary Table 7. Distribution of death and recurrence according to antibiotic intake.**

| | **Localized colon cancer** n=17 735 | | **Advanced colon cancer** n=5 805 | | **Localized rectum/junction cancer** n=5 097 | | **Advanced rectum/junction cancer** n=6 859 | | **Total** n=35 496 | |
|---|---|---|---|---|---|---|---|---|---|---|
| | n | % | n | % | n | % | n | % | n | % |
| **No ATB intake** | 4 094 | | 964 | | 1 240 | | 1 281 | | 7 577 | |
| no recurrence | 2 891 | 70.6 | 623 | 64.6 | 858 | 69.2 | 609 | 47.5 | 4 979 | 65.7 |
| recurrence or death | 1 203 | 29.4 | 341 | 35.4 | 382 | 30.8 | 672 | 52.5 | 2 598 | 34.3 |
| recurrence | 764 | 18.7 | 320 | 33.2 | 288 | 23.2 | 624 | 48.7 | 1 996 | 26.3 |
| death | 439 | 10.7 | 21 | 2.2 | 94 | 7.6 | 48 | 3.7 | 602 | 7.9 |
| of which death at 2 months | 254 | 6.2 | 4 | 0.4 | 48 | 3.9 | 34 | 2.7 | 340 | 4.5 |
| **Only hospital** | 1 862 | | 418 | | 465 | | 585 | | 3 330 | |
| no recurrence | 854 | 45.9 | 206 | 49.3 | 187 | 40.2 | 217 | 37.1 | 1 464 | 44.0 |
| recurrence or death | 1 008 | 54.1 | 212 | 50.7 | 278 | 59.8 | 368 | 62.9 | 1 866 | 56.0 |
| recurrence | 446 | 24.0 | 179 | 42.8 | 169 | 36.3 | 292 | 49.9 | 1 086 | 32.6 |
| death | 562 | 30.2 | 33 | 7.9 | 109 | 23.4 | 76 | 13.0 | 780 | 23.4 |
| of which death at 2 months | 368 | 19.8 | 11 | 2.6 | 71 | 15.3 | 38 | 6.5 | 488 | 14.7 |
| **Only city pharmacy** | 7 025 | | 2 692 | | 2 124 | | 2 752 | | 14 593 | |
| no recurrence | 5 298 | 75.4 | 1 702 | 63.2 | 1 585 | 74.6 | 1 456 | 52.9 | 10 041 | 68.8 |
| recurrence or death | 1 727 | 24.6 | 990 | 36.8 | 539 | 25.4 | 1 296 | 47.1 | 4 552 | 31.2 |
| recurrence | 1 249 | 17.8 | 940 | 34.9 | 455 | 21.4 | 1 236 | 44.9 | 3 880 | 26.6 |
| death | 478 | 6.8 | 50 | 1.9 | 84 | 4.0 | 60 | 2.2 | 672 | 4.6 |
| of which death at 2 months | 191 | 2.7 | 10 | 0.4 | 35 | 1.6 | 19 | 0.7 | 255 | 1.7 |
| **Hospital and city pharmacy** | 4 754 | | 1 733 | | 1 268 | | 2 241 | | 9 996 | |
| no recurrence | 2 930 | 61.6 | 925 | 53.4 | 724 | 57.1 | 928 | 41.4 | 5 507 | 55.1 |
| recurrence or death | 1 824 | 38.4 | 808 | 46.6 | 544 | 42.9 | 1 313 | 58.6 | 4 489 | 44.9 |
| recurrence | 1 094 | 23.0 | 748 | 43.2 | 415 | 32.7 | 1 221 | 54.5 | 3 478 | 34.8 |
| death | 730 | 15.4 | 60 | 3.5 | 129 | 10.2 | 92 | 4.1 | 1 011 | 10.1 |
| of which death at 2 months | 312 | 6.6 | 11 | 0.6 | 52 | 4.1 | 27 | 1.2 | 402 | 4.0 |

**Supplementary Table 8. Distribution of death and recurrence according to antibiotic intake 2 months after resection.**

| | No hospitalization for infection n=22 199 | | Hospitalization for infection n=13 297 | | |
|---|---|---|---|---|---|
| | n | % | n | % | p-value |
| **Sex:** | | | | | <0,0001 |
| Male | 11 310 | 51.0 | 7 250 | 54.4 | |
| Female | 10 860 | 49.0 | 6 076 | 45.6 | |
| **Age, years:** | | | | | <0,0001 |
| 18-49 | 1 536 | 6.9 | 817 | 6.1 | |
| 50-69 | 9 878 | 44.6 | 4 935 | 37.0 | |
| 70-79 | 5 697 | 25.7 | 3 467 | 26.0 | |
| ≥ 80 | 5 059 | 22.8 | 4 107 | 30.8 | |
| **Malnutrition:** | | | | | <0,0001 |
| No | 20 278 | 91.5 | 10 968 | 82.3 | |
| Yes | 1 892 | 8.5 | 2 358 | 17.7 | |
| **Charlson comorbidities:** | | | | | <0,0001 |
| 0 | 14 878 | 67.1 | 7 834 | 58.8 | |
| 1-2 | 5 645 | 25.5 | 3 781 | 28.4 | |
| ≥ 3 | 1 647 | 7.4 | 1 711 | 12.8 | |
| **Length of stay in intensive care unit, days:** | | | | | <0,0001 |
| 0-2 | 21 286 | 96.0 | 11 551 | 86.7 | |
| 2-7 | 651 | 2.9 | 824 | 6.2 | |
| > 7 | 233 | 1.1 | 951 | 7.1 | |

### Supplementary Table 9. Multivariate analyses for 3-year DFS of patients with resected colorectal cancer according to antibiotic and chemotherapy exposure.

#### 9A-Localized colon cancer patients (n=11,140)

| Analysis of Maximum Likelihood Estimates | | | | | |
|---|---|---|---|---|---|
| **Parameter** | | **Pr > ChiSq** | **Hazard Ratio** | **95% Hazard Ratio Confidence Limits** | |
| Sex | Male *vs* Female | <.0001 | 1 | 1.111 | 1.289 |
| Age | 50 to 69 *vs* <50 | 0.2501 | 1.135 | 0.915 | 1.409 |
| | 70-79 *vs* <50 | 0.0009 | 1.446 | 1.162 | 1.798 |
| | ≥ 80 *vs* <50 | <.0001 | 2.498 | 2.018 | 3.092 |
| Charlson | 1 à 2 *vs* 0 | <.0001 | 1.223 | 1.125 | 1.330 |
| | ≥ 3 *vs* 0 | <.0001 | 1.700 | 1.527 | 1.893 |
| Malnutrition | Yes *vs* No | <.0001 | 1.526 | 1.373 | 1.697 |
| ICU stay | > 2 to 7 days *vs* 0-2 days | <.0001 | 1.480 | 1.231 | 1.779 |
| | > 7 days *vs* 0-2 days | <.0001 | 2.179 | 1.702 | 2.789 |
| *Pre resection ATB exposure* | | | | | |
| ATB as outpatient | Yes *vs* No | 0.8253 | 1.009 | 0.932 | 1.093 |
| *Post resection ATB exposure* | | | | | |
| **ATB as outpatient** | **Yes *vs* No** | **0.0169** | **1.105** | **1.018** | **1.199** |

| Analysis of Maximum Likelihood Estimates | | | | | |
|---|---|---|---|---|---|
| **Parameter** | | **Pr > ChiSq** | **Hazard Ratio** | **95% Hazard Ratio Confidence Limits** | |
| Sex | Male *vs* Female | <.0001 | 1.194 | 1.108 | 1.287 |
| Age | 50 to 69 *vs* <50 | 0.2476 | 1.136 | 0.915 | 1.411 |
| | 70-79 *vs* <50 | 0.0009 | 1.450 | 1.165 | 1.803 |
| | ≥ 80 *vs* <50 | <.0001 | 2.501 | 2.020 | 3.096 |
| Charlson | 1 à 2 *vs* 0 | <.0001 | 1.221 | 1.123 | 1.328 |
| | ≥ 3 *vs* 0 | <.0001 | 1.696 | 1.523 | 1.889 |
| Malnutrition | Yes *vs* No | <.0001 | 1.522 | 1.368 | 1.692 |
| ICU stay | > 2 to 7 days *vs* 0-2 days | <.0001 | 1.470 | 1.223 | 1.768 |
| | > 7 days *vs* 0-2 days | <.0001 | 2.145 | 1.675 | 2.746 |
| *Pre resection ATB exposure* | | | | | |
| ATB as outpatient | Yes *vs* No | 0.9421 | 1.003 | 0.926 | 1.086 |
| *Post resection ATB exposure* | | | | | |
| C3G | Yes *vs* No | 0.7560 | 0.977 | 0.843 | 1.132 |
| Antifungal – Others | Yes *vs* No | 0.1525 | 1.189 | 0.938 | 1.506 |
| **Antifungal – Azoles** | **Yes *vs* No** | **0.0050** | **1.446** | **1.118** | **1.870** |
| Beta-lactamines – Others | Yes *vs* No | 0.5139 | 1.069 | 0.876 | 1.304 |
| Penicillin A | Yes *vs* No | 0.6623 | 0.976 | 0.876 | 1.088 |
| **Penicillin A + beta-lactamase inhibitor** | **Yes *vs* No** | **0.0127** | **1.149** | **1.030** | **1.282** |
| Macrolides | Yes *vs* No | 0.2413 | 0.913 | 0.785 | 1.063 |
| Metronidazole based-antibiotherapies | Yes *vs* No | 0.7275 | 1.026 | 0.887 | 1.188 |
| **Quinolones** | **Yes *vs* No** | **0.0087** | **1.160** | **1.038** | **1.297** |
| Streptogramins | Yes *vs* No | 0.1000 | 1.183 | 0.968 | 1.444 |
| Other ATB including cyclins | Yes *vs* No | 0.9900 | 0.999 | 0.856 | 1.167 |

### 9B-Advanced colon cancer patients (n=5,805)

| Analysis of Maximum Likelihood Estimates | | | | | |
|---|---|---|---|---|---|
| **Parameter** | | **Pr > ChiSq** | **Hazard Ratio** | **95% Hazard Ratio Confidence Limits** | |
| Sex | Male *vs* Female | 0.0010 | 1.203 | 1.078 | 1.343 |
| Age | 50 to 69 *vs* <50 | 0.0616 | 1.221 | 0.990 | 1.506 |
| | 70-79 *vs* <50 | 0.0498 | 1.248 | 1.000 | 1.558 |
| | ≥ 80 *vs* <50 | 0.0002 | 1.600 | 1.246 | 2.055 |
| Charlson | 1 à 2 *vs* 0 | 0.0592 | 1.129 | 0.995 | 1.281 |
| | ≥ 3 *vs* 0 | 0.9492 | 0.992 | 0.775 | 1.270 |
| Malnutrition | Yes *vs* No | 0.0110 | 1.256 | 1.054 | 1.496 |
| ICU stay | > 2 to 7 days *vs* 0-2 days | 0.9385 | 1.014 | 0.720 | 1.427 |
| | > 7 days *vs* 0-2 days | 0.0236 | 1.769 | 1.080 | 2.899 |
| *Pre resection ATB exposure* | | | | | |
| ATB as outpatient | Yes *vs* No | 0.5452 | 1.036 | 0.924 | 1.163 |
| *Post resection ATB exposure* | | | | | |
| **ATB as outpatient** | **Yes *vs* No** | **<.0001** | **1.280** | **1.134** | **1.446** |

| Analysis of Maximum Likelihood Estimates | | | | | |
|---|---|---|---|---|---|
| **Parameter** | | **Pr > ChiSq** | **Hazard Ratio** | **95% Hazard Ratio Confidence Limits** | |
| Sex | Male *vs* Female | 0.0011 | 1.200 | 1.075 | 1.340 |
| Age | 50 to 69 *vs* <50 | 0.0900 | 1.199 | 0.972 | 1.479 |
| | 70-79 *vs* <50 | 0.0499 | 1.249 | 1.000 | 1.560 |
| | ≥ 80 *vs* <50 | 0.0002 | 1.619 | 1.259 | 2.081 |
| Charlson | 1 à 2 *vs* 0 | 0.0426 | 1.139 | 1.004 | 1.293 |
| | ≥ 3 *vs* 0 | 0.4683 | 0.912 | 0.710 | 1.171 |
| Malnutrition | Yes *vs* No | 0.0067 | 1.275 | 1.070 | 1.519 |
| ICU stay | > 2 to 7 days *vs* 0-2 days | 0.9899 | 1.002 | 0.712 | 1.412 |
| | > 7 days *vs* 0-2 days | 0.0186 | 1.810 | 1.104 | 2.968 |
| *Pre resection ATB exposure* | | | | | |
| ATB as outpatient | ATB/no neoadjuvant *vs* no ATB/no neoadjuvant | 0.7760 | 0.983 | 0.870 | 1.109 |
| Timing of chemotherapy and antibiotherapy | ATB outside neoadjuvant *vs* Neoadjuvant/no ATB | 0.6663 | 1.169 | 0.575 | 2.375 |
| | ATB during neoadjuvant *vs* Neoadjuvant/no ATB | 0.6971 | 1.091 | 0.703 | 1.694 |
| *Post resection ATB exposure* | | | | | |
| **ATB outside chemotherapy** | **Yes *vs* No** | **0.0361** | **1.213** | **1.013** | **1.453** |
| **ATB during chemotherapy** | **Yes *vs* No** | **<.0001** | **1.317** | **1.162** | **1.493** |

| Analysis of Maximum Likelihood Estimates | | | | | |
|---|---|---|---|---|---|
| **Parameter** | | **Pr > ChiSq** | **Hazard Ratio** | **95% Hazard Ratio Confidence Limits** | |
| Sex | Male *vs* Female | 0.0001 | 1.241 | 1.111 | 1.387 |
| Age | 50 to 69 *vs* <50 | 0.0839 | 1.203 | 0.976 | 1.484 |
| | 70-79 *vs* <50 | 0.0639 | 1.234 | 0.988 | 1.541 |
| | ≥ 80 *vs* <50 | 0.0002 | 1.615 | 1.255 | 2.077 |
| Charlson | 1 à 2 *vs* 0 | 0.0292 | 1.151 | 1.014 | 1.307 |

| | | | | | |
|---|---|---|---|---|---|
| | ≥ 3 *vs* 0 | 0.6057 | 0.936 | 0.728 | 1.203 |
| Malnutrition | Yes *vs* No | 0.0045 | 1.291 | 1.082 | 1.539 |
| ICU stay | > 2 to 7 days *vs* 0-2 days | 0.8561 | 0.969 | 0.687 | 1.365 |
| | > 7 days *vs* 0-2 days | 0.0107 | 1.908 | 1.161 | 3.133 |
| *Pre resection ATB exposure* | | | | | |
| ATB as outpatient | ATB/no neoadjuvant *vs* no ATB/no neoadjuvant | 0.6138 | 0.969 | 0.857 | 1.095 |
| Timing of chemotherapy and antibiotherapy | ATB outside neoadjuvant *vs* Neoadjuvant/no ATB | 0.7539 | 1.121 | 0.550 | 2.284 |
| | ATB during neoadjuvant *vs* Neoadjuvant/no ATB | 0.9516 | 0.986 | 0.632 | 1.539 |
| *Post resection ATB exposure* | | | | | |
| **C3G** | **Yes *vs* No** | **0.0290** | **1.252** | **1.023** | **1.531** |
| **Antifungal – Others** | **Yes *vs* No** | **0.0056** | **1.217** | **1.059** | **1.398** |
| Antifungal – Azoles | Yes *vs* No | 0.4257 | 0.926 | 0.768 | 1.118 |
| Beta-lactamines – Others | Yes *vs* No | 0.2256 | 0.831 | 0.615 | 1.121 |
| Penicillin A | Yes *vs* No | 0.2399 | 1.088 | 0.945 | 1.252 |
| Penicillin A + beta-lactamase inhibitor | Yes *vs* No | 0.7275 | 1.024 | 0.895 | 1.173 |
| Macrolides | Yes *vs* No | 0.8848 | 0.985 | 0.803 | 1.209 |
| Metronidazole based-antibiotherapies | Yes *vs* No | 0.6575 | 1.044 | 0.864 | 1.261 |
| **Quinolones** | **Yes *vs* No** | **0.0411** | **1.157** | **1.006** | **1.331** |
| Streptogramins | Yes *vs* No | 0.5780 | 1.079 | 0.826 | 1.409 |
| **Other ATB including cyclins** | **Yes *vs* No** | **<.0001** | **1.553** | **1.299** | **1.856** |

### 9C-Localized rectum/junction cancer (N=3,364)

| Analysis of Maximum Likelihood Estimates | | | | | |
|---|---|---|---|---|---|
| **Parameter** | | **Pr > ChiSq** | **Hazard Ratio** | **95% Hazard Ratio Confidence Limits** | |
| Sex | Male *vs* Female | <.0001 | 1.425 | 1.248 | 1.628 |
| Age | 50 to 69 *vs* <50 | 0.5045 | 1.125 | 0.797 | 1.588 |
| | 70-79 *vs* <50 | 0.0076 | 1.617 | 1.136 | 2.301 |
| | ≥ 80 *vs* <50 | <.0001 | 2.268 | 1.596 | 3.224 |
| Charlson | 1 à 2 *vs* 0 | 0.1901 | 1.107 | 0.951 | 1.288 |
| | ≥ 3 *vs* 0 | 0.0126 | 1.340 | 1.065 | 1.686 |
| Malnutrition | Yes *vs* No | 0.0015 | 1.448 | 1.152 | 1.820 |
| ICU stay | > 2 to 7 days *vs* 0-2 days | 0.4724 | 1.133 | 0.807 | 1.590 |
| | > 7 days *vs* 0-2 days | 0.2320 | 1.316 | 0.839 | 2.064 |
| *Pre resection ATB exposure* | | | | | |
| ATB as outpatient | Yes *vs* No | 0.8955 | 1.010 | 0.875 | 1.164 |
| *Post resection ATB exposure* | | | | | |
| ATB as outpatient | Yes *vs* No | 0.5181 | 1.049 | 0.908 | 1.212 |

| Analysis of Maximum Likelihood Estimates | | | |
|---|---|---|---|
| **Parameter** | **Pr > ChiSq** | | **95% Hazard Ratio** |

| | | | Hazard Ratio | Confidence Limits | |
|---|---|---|---|---|---|
| Sex | Male *vs* Female | <.0001 | 1.442 | 1.261 | 1.650 |
| Age | 50 to 69 *vs* <50 | 0.4666 | 1.137 | 0.805 | 1.605 |
| | 70-79 *vs* <50 | 0.0065 | 1.632 | 1.147 | 2.323 |
| | ≥ 80 *vs* <50 | <.0001 | 2.301 | 1.618 | 3.271 |
| Charlson | 1 à 2 *vs* 0 | 0.2987 | 1.084 | 0.931 | 1.263 |
| | ≥ 3 *vs* 0 | 0.0146 | 1.333 | 1.058 | 1.679 |
| Malnutrition | Yes *vs* No | 0.0012 | 1.462 | 1.163 | 1.839 |
| ICU stay | > 2 to 7 days *vs* 0-2 days | 0.5158 | 1.119 | 0.797 | 1.572 |
| | > 7 days *vs* 0-2 days | 0.2444 | 1.308 | 0.832 | 2.057 |
| *Pre resection ATB exposure* | | | | | |
| ATB as outpatient | Yes *vs* No | 0.6528 | 0.967 | 0.838 | 1.117 |
| *Post resection ATB exposure* | | | | | |
| C3G | Yes *vs* No | 0.6601 | 1.070 | 0.792 | 1.445 |
| Antifungal – Others | Yes *vs* No | 0.4697 | 1.213 | 0.718 | 2.050 |
| Antifungal – Azoles | Yes *vs* No | 0.7559 | 1.098 | 0.610 | 1.973 |
| Beta-lactamines – Others | Yes *vs* No | 0.4107 | 0.833 | 0.539 | 1.288 |
| Penicillin A | Yes *vs* No | 0.7019 | 1.039 | 0.854 | 1.264 |
| **Penicillin A + beta-lactamase inhibitor** | **Yes *vs* No** | **0.0223** | **1.255** | **1.033** | **1.525** |
| Macrolides | Yes *vs* No | 0.9791 | 1.003 | 0.773 | 1.303 |
| Metronidazole based-antibiotherapies | Yes *vs* No | 0.7444 | 0.962 | 0.760 | 1.217 |
| **Quinolones** | **Yes *vs* No** | **0.0223** | **1.258** | **1.033** | **1.532** |
| Streptogramins | Yes *vs* No | 0.9792 | 1.005 | 0.700 | 1.443 |
| Other ATB including cyclins | Yes *vs* No | 0.2360 | 1.174 | 0.900 | 1.531 |

### 9D-Advanced rectum/junction cancer patients (N=4,034)

| Analysis of Maximum Likelihood Estimates | | | | | |
|---|---|---|---|---|---|
| **Parameter** | | **Pr > ChiSq** | **Hazard Ratio** | **95% Hazard Ratio Confidence Limits** | |
| Sex | Male *vs* Female | 0.0021 | 1.155 | 1.054 | 1.266 |
| Age | 50 to 69 *vs* <50 | 0.7527 | 0.975 | 0.834 | 1.140 |
| | 70-79 *vs* <50 | 0.8535 | 0.984 | 0.828 | 1.169 |
| | ≥ 80 *vs* <50 | 0.0348 | 1.238 | 1.015 | 1.509 |
| Charlson | 1 à 2 *vs* 0 | 0.7465 | 1.018 | 0.913 | 1.135 |
| | ≥ 3 *vs* 0 | 0.8708 | 1.018 | 0.823 | 1.259 |
| Malnutrition | Yes *vs* No | 0.0076 | 1.215 | 1.053 | 1.402 |
| ICU stay | > 2 to 7 days *vs* 0-2 days | 0.1464 | 0.838 | 0.660 | 1.064 |
| | > 7 days *vs* 0-2 days | 0.8236 | 0.950 | 0.603 | 1.495 |
| *Pre resection ATB exposure* | | | | | |
| ATB as outpatient | Yes *vs* No | 0.6389 | 1.023 | 0.930 | 1.125 |
| *Post resection ATB exposure* | | | | | |
| ATB as outpatient | Yes *vs* No | 0.1004 | 1.083 | 0.985 | 1.191 |
| **Analysis of Maximum Likelihood Estimates** | | | | | |
| **Parameter** | | **Pr > ChiSq** | **Hazard Ratio** | **95% Hazard Ratio Confidence Limits** | |

| | | | | | |
|---|---|---|---|---|---|
| Sex | Male *vs* Female | 0.0072 | 1.135 | 1.035 | 1.244 |
| Age | 50 to 69 *vs* <50 | 0.9004 | 0.990 | 0.847 | 1.158 |
| | 70-79 *vs* <50 | 0.7987 | 1.023 | 0.860 | 1.216 |
| | ≥ 80 *vs* <50 | 0.0005 | 1.423 | 1.165 | 1.739 |
| Charlson | 1 à 2 *vs* 0 | 0.3990 | 1.048 | 0.940 | 1.168 |
| | ≥ 3 *vs* 0 | 0.7149 | 1.040 | 0.841 | 1.287 |
| Malnutrition | Yes *vs* No | 0.0178 | 1.189 | 1.030 | 1.371 |
| ICU stay | > 2 to 7 days *vs* 0-2 days | 0.1287 | 0.831 | 0.655 | 1.055 |
| | > 7 days *vs* 0-2 days | 0.9471 | 0.985 | 0.626 | 1.550 |
| *Pre resection ATB exposure* | | | | | |
| ATB as outpatient | ATB/no neoadjuvant *vs* no ATB/no neoadjuvant | 0.5251 | 1.057 | 0.891 | 1.255 |
| Timing of chemotherapy and antibiotherapy | ATB outside neoadjuvant *vs* Neoadjuvant/no ATB | 0.1612 | 0.895 | 0.765 | 1.045 |
| | ATB during neoadjuvant *vs* Neoadjuvant/no ATB | 0.1880 | 1.091 | 0.958 | 1.242 |
| *Post resection ATB exposure* | | | | | |
| ATB outside chemotherapy | Yes *vs* No | 0.1082 | 0.910 | 0.811 | 1.021 |
| **ATB during chemotherapy** | **Yes *vs* No** | **<.0001** | **1.412** | **1.259** | **1.584** |

**Analysis of Maximum Likelihood Estimates**

| Parameter | | Pr > ChiSq | Hazard Ratio | 95% Hazard Ratio Confidence Limits | |
|---|---|---|---|---|---|
| Sex | Male *vs* Female | 0.0028 | 1.151 | 1.050 | 1.263 |
| Age | 50 to 69 *vs* <50 | 0.8441 | 0.984 | 0.841 | 1.152 |
| | 70-79 *vs* <50 | 0.9839 | 1.002 | 0.842 | 1.192 |
| | ≥ 80 *vs* <50 | 0.0042 | 1.339 | 1.097 | 1.635 |
| Charlson | 1 à 2 *vs* 0 | 0.5948 | 1.030 | 0.924 | 1.149 |
| | ≥ 3 *vs* 0 | 0.6795 | 1.046 | 0.845 | 1.294 |
| Malnutrition | Yes *vs* No | 0.0137 | 1.197 | 1.038 | 1.382 |
| ICU stay | > 2 to 7 days *vs* 0-2 days | 0.1006 | 0.818 | 0.644 | 1.040 |
| | > 7 days *vs* 0-2 days | 0.8184 | 0.948 | 0.602 | 1.493 |
| *Pre resection ATB exposure* | | | | | |
| ATB as outpatient | ATB/no neoadjuvant *vs* no ATB/no neoadjuvant | 0.9659 | 0.996 | 0.837 | 1.186 |
| Timing of chemotherapy and antibiotherapy | **ATB outside neoadjuvant *vs* Neoadjuvant/no ATB** | **0.0295** | **0.840** | **0.719** | **0.983** |
| | ATB during neoadjuvant *vs* Neoadjuvant/no ATB | 0.5389 | 1.042 | 0.914 | 1.187 |
| *Post resection ATB exposure* | | | | | |
| C3G | Yes *vs* No | 0.0519 | 1.206 | 0.998 | 1.458 |
| Antifungal – Others | Yes *vs* No | 0.4835 | 1.061 | 0.899 | 1.252 |
| Antifungal – Azoles | Yes *vs* No | 0.0768 | 1.182 | 0.982 | 1.422 |
| Beta-lactamines – Others | Yes *vs* No | 0.2122 | 1.190 | 0.905 | 1.564 |
| Penicillin A | Yes *vs* No | 0.8288 | 1.015 | 0.885 | 1.164 |
| Penicillin A + beta-lactamase inhibitor | Yes *vs* No | 0.0607 | 1.125 | 0.995 | 1.271 |
| Macrolides | Yes *vs* No | 0.5720 | 0.943 | 0.769 | 1.156 |
| **Metronidazole based-antibiotherapies** | **Yes *vs* No** | **0.0132** | **1.204** | **1.040** | **1.395** |
| Quinolones | Yes *vs* No | 0.8391 | 1.012 | 0.900 | 1.138 |
| Streptogramins | Yes *vs* No | 0.3152 | 1.134 | 0.887 | 1.450 |
| **Other ATB including cyclins** | **Yes *vs* No** | **<.0001** | **1.402** | **1.197** | **1.643** |

**Supplementary Figure 1. Distribution of the number of chemotherapy cycles according to antibiotic intake in the groups of patients receiving at least one adjuvant chemotherapy cycle (N=9,205).**

N.B: In France, when reimbursing the cost of chemotherapy, a 2-day treatment such as LV5FU2 or FOLFOX is considered to be 2 cycles if the patient goes to the hospital on both days (D1 and D2). This can explain why some patients receive more than the usual 12 cycles of chemotherapy in the adjuvant setting.

| | No antibiotic | Antibiotic |
|---|---|---|
| Min | 1 | 1 |
| $Q_1$ | 11 | 11 |
| Median | 12 | 12 |
| $Q_3$ | 22 | 24 |
| Max | 155 | 203 |

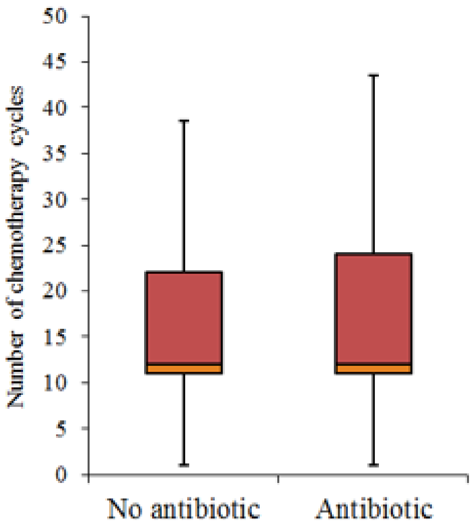

**Supplementary Figure 2. Three-year disease-free survival of resected colorectal cancer patients according to the primitive location and stage.**

Univariate 3-year disease-free survival assessing the impact of the primitive location and stage (N=35,496).

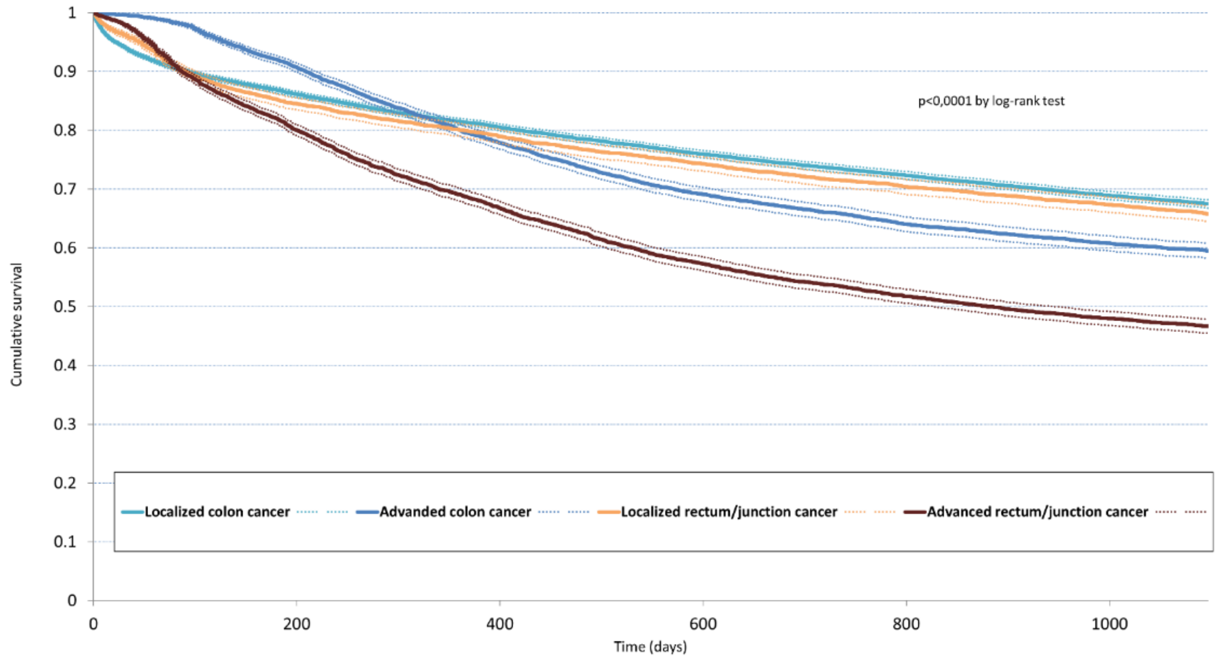
